# Modelling a potential zoonotic spillover event of H5N1 influenza

**DOI:** 10.1101/2025.04.28.25326570

**Authors:** Philip Cherian, Gautam I. Menon

**Affiliations:** Department of Physics, Ashoka University, Sonepat, Haryana, India; Niels Bohr Institute, University of Copenhagen, Copenhagen, Denmark; Department of Biology, Trivedi School of Biological Sciences, Ashoka University, Sonepat, Haryana, India; Center for Climate Change and Sustainability, Ashoka University, Sonepat, Haryana, India

## Abstract

**Background:** Highly Pathogenic Avian Influenza (HPAI) is a prominent candidate for a future human pandemic arising from a zoonotic spillover event. Its best-known subtype is H5N1, with South-or South-East Asia a likely location for an initial outbreak. Such an outbreak would be initiated through a primary event of bird-to-human infection, followed by sustained human-to-human transmission. Early interventions require the extraction, integration and interpretation of epidemiological information from the limited and noisy case data available at outbreak onset.

**Methods:** We studied the implications of a potential zoonotic spillover of H5N1 influenza into humans. Our simulations used *BharatSim*, an agent-based model framework designed primarily for the population of India, but which can be tuned easily for others. We considered a synthetic population representing primary contacts in an outbreak site with infected birds. These primary contacts transfer infections to secondary (household) contacts, from where the infection spreads further. We simulate outbreak scenarios in farm as well as wet-market settings, accounting for the network structure of human contacts and the stochasticity of the infection process. We further simulated multiple interventions, including bird-culling, quarantines, and vaccinations.

**Results:** We show how limited, noisy data for primary and secondary infections can be used to estimate epidemiological transmission parameters, such as the basic reproductive ratio *R*_0_ from other metrics like the secondary attack risk, in realistic social interaction settings. We describe the impact of early interventions (bird-culling, quarantines, and vaccination), taken together or separately, in slowing or terminating the outbreak.

**Conclusions:** An individual-based model allows for the most granular description of the bird-human spillover and subsequent human-to-human transmission for the case of H5N1. Such models can be contextualised to individual communities across varied geographies, given representative contact networks. We show how such models allow for the systematic real-time exploration of policy measures that could constrain disease-spread, as well as guide a better understanding of disease epidemiology for an emerging infectious disease.

## 1. Introduction

A likely scenario for a future Disease X [1, 2] associates it with a viral infection in humans, initiated through a zoonotic spillover event. Such an infection would transmit efficiently via a respiratory route. A strong candidate for such a disease is highly pathogenic avian influenza (HPAI), commonly called “bird-flu” [3–5].

The most prominent example of HPAI is H5N1 influenza, a disease very largely confined to birds, but with a demonstrated ability to infect both terrestrial and marine mammals [4, 6–8]. There have, so far, only been isolated reports of bird-to-human transmission, either directly or via household pets. There are no confirmed reports of human-to-human transmission [9]. However, in the few known cases of human infection, case fatality rates of more than 30% have been suggested [10, 11], pointing to the importance of planning for a future outbreak.

The initial stages of any outbreak of a novel disease are inevitably marked by uncertainty as to the epidemiological parameters that characterise its spread [12]. The epidemiology here must account for two bottlenecks [13]. The first is the ability of the virus to cause infections in human hosts, while the second describes the barriers to subsequent human-to-human transmission. Once the initial threshold of viral adaptability is breached, the potential to cause a pandemic depends on the efficiency of human-to-human transmission.

The usefulness of models is that they allow for the reconstruction of potential disease trajectories from which, by comparison, such epidemiological parameters and their consequences can be inferred. A variety of disease models can be used to describe an outbreak. They typically differ in the scale of their description. The model may be defined at the level of groups or categories of individuals (susceptible or infected, as in conventional compartmental models) or at the level of single individuals, where individual-level variation and networks of potential contacts leading to infection must be accounted for. A number of models have been used to describe avian influenza and its transmission both between birds [14–18] and into mammals [4, 19–22].

Here, since the early stages of an outbreak are marked by a small number of infections, each of them occurring through a stochastic infection event, any useful analysis must account for multiple alternative disease trajectories describing transmission between individuals, a task for which agent-based simulations are best suited. In this paper, we simulate potential scenarios of an H5N1 outbreak in humans.

While zoonotic spillover and human-to-human transmission have been extensively studied in the context of pandemic preparedness, several models in the existing literature have already attempted to capture key aspects of this two-step process. For example, van Boven et al. (2007) developed a household transmission model to detect the emerging transmissibility of avian influenza within human households, highlighting the challenges of early detection in cluster-based data [23]. Yang et al. (2007) similarly proposed statistical methods to infer human-to-human transmissibility of H5N1 using outbreak data [24], while Iwami et al. (2007) presented a deterministic compartmental model of avian-to-human influenza transmission to explore epidemic thresholds [25]. Other work has taken more inference-driven approaches: Bettencourt and Ribeiro (2008) introduced a real-time Bayesian framework for estimating epidemic potential [26], and Lo Iacono et al. (2016) proposed a unified framework that integrates zoonotic spillover and subsequent spread in humans [27]. More recently, Saldaña et al. (2024) provided a detailed overview of modelling frameworks for spillover dynamics of emerging pathogens, emphasizing ecological and evolutionary processes underpinning transmission [28].

Here, we explicitly model a two-step zoonotic spillover scenario in a realistic population that is tailored to a low-and middle-income country (LMIC) setting. Our work goes beyond prior work in that it explicitly accounts for contact networks and family structures that are appropriate for a developing country in South Asia, a probable source of an HPAI pandemic. Our simulation-based framework enables the incorporation of heterogeneity in individual contact patterns, in infection progression, and intervention response, contributing insights that complement and advance the existing literature.

Our simulations use *BharatSim*, an agent-based simulation framework developed for India [29]. Agent-based models are the most granular of models since they are defined at the level of single individuals and the networks of their interactions. While BharatSim’s capabilities extend to simulations of up to 50 million individual agents, a much smaller population of approximately 10,000 agents is sufficient for us to be able to address the early stages of an outbreak.

BharatSim uses a synthetic population of considerable complexity, incorporating social and economic information obtained from combining a variety of surveys and census information using refined AI/ML techniques [29, 30]. Much of this complexity is irrelevant to the questions we ask here, so we choose simpler representations, arguing that these should generalize across multiple LMIC geographies as well. Our simulations work with networks of homes, workplaces (including schools) and the interactions between primary, secondary and tertiary contacts across them. We describe the construction of the synthetic population in Section 2.2.

The ability to describe a range of different disease trajectories while also potentially accommodating individual-level heterogeneity is a feature of the simulation methodology described in this paper. We use results from these simulations to extract information about parameters governing epidemiological transmission, such as the basic reproductive ratio and the secondary attack risk, across a range of scenarios. Our simulation methods allow us to explore targeted interventions and synergies between them, for example isolating and quarantining populations vulnerable to infection [31, 32].

## 2 Methods

Our simulations of a potential H5N1 epidemic originating in a spillover to humans use an agent-based approach for disease-spread in the human population. This disease is seeded through infected birds within a single location (the outbreak site).

### 2.1 Model

A schematic of our model, implemented within BharatSim, is shown in Fig. 1(A). This describes an outbreak site with infected birds and a set of primary contacts. These primary contacts have secondary contacts at their homes.

**Fig. 1:**
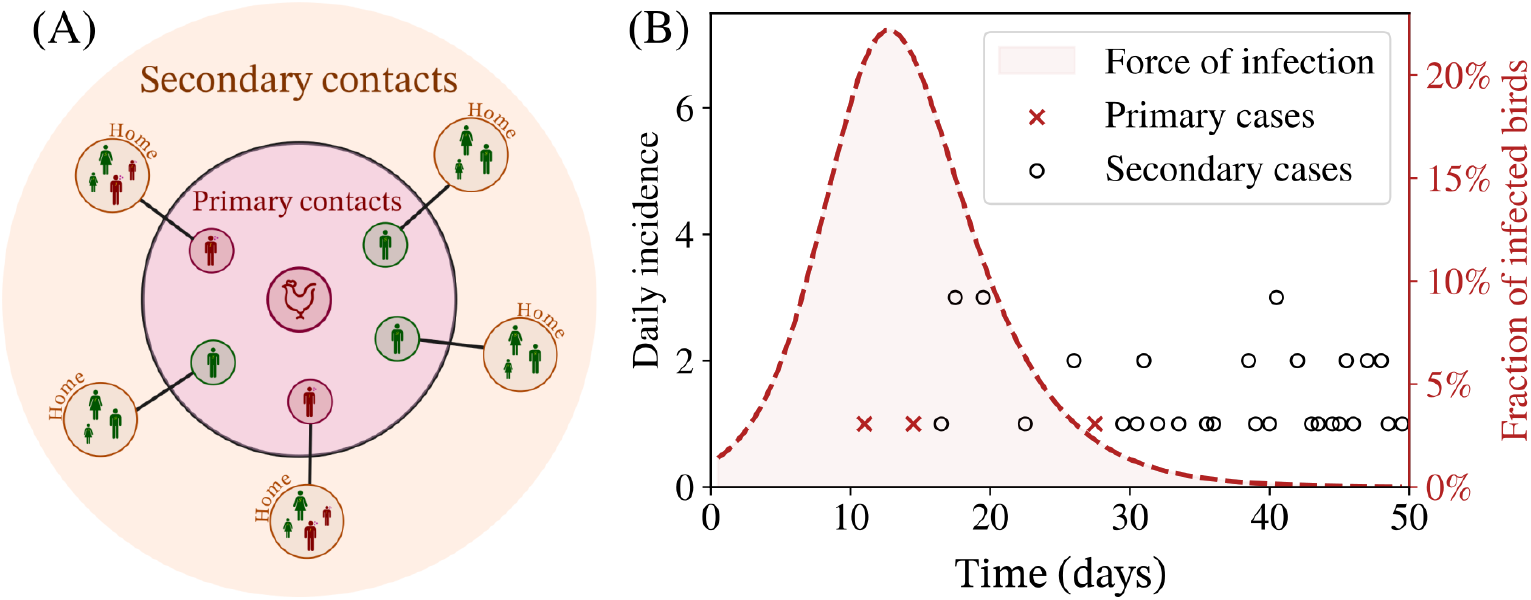
(A) Schematic illustrating primary and secondary contacts of infected birds associated with the outbreak site. For example, in the farm-outbreak scenario, primary contacts are assumed to be the poultry farmers, who can then pass the infection to their household (secondary) contacts at home. Infected agents are shown in red while susceptible agents are green. (B) A representative time-series of daily incidence in the case of a spillover event for both primary (crosses) and secondary cases (circles) at the onset of the outbreak. Also shown is the fraction of infected birds in outbreak site (assuming a farm-outbreak), directly proportional to the force of infection.

The individuals in our model are part of larger networks. They move between different locations during a single day, reconfiguring their contact networks. BharatSim assumes that any individual above the age of 18 works at a defined ‘workplace’, while the remaining are children who go to schools. Agents spend 12 hours at their homes, interacting with their household contacts. The remaining 12 hours are spent at their workplaces or schools, where they interact with a different set of contacts. A small fraction of agents in the population are homebound, spending all their time at home. One specific workplace is designated as the outbreak site, from where the infection is initiated, with individuals working at this location being primary contacts.

### 2.2 Generating a synthetic population

The BharatSim framework allows users the flexibility to generate a synthetic population of a specific region, by applying statistical methods and machine learning algorithms to survey data from multiple sources, including the Census of India, the India Human Development Survey, the National Sample Survey, and the Gridded Population of the World. This synthetic population defines individual agents with multiple attributes, among them age, gender, home and work locations, pre-existing health conditions, and socio-economic and employment status.

For concreteness, in this study we restrict ourselves to a region of study that comprises a single small village in the district of Namakkal in the state of Tamil Nadu in India. The Namakkal district houses more than 1600 poultry farms that rear over 70 million chickens. Producing over 60 million eggs in a day, this district supplies most of the eggs for the Indian states of Tamil Nadu, Kerala, and Karnataka [33]. We model a village with a population of 9,667 individuals, and use the synthetic population pipeline described in detail in Refs. [29, 30] to generate a population that harmonises aggregate data from the 2011 Census and the National Sample Survey along with micro-data from the India Human Development Survey. The households and workplaces are distributed by gridded population density from the Gridded Population of the World.

A single workplace is designated as the outbreak site in our simulations. This site comprises 165 farmers, with 476 secondary contacts. The network structure of homes, workplaces, and schools remains fixed through our different simulations. A small fraction of our agents (5%) are assumed to be home-bound, meaning that they do not travel between homes and workplaces. Further information about the population can be found in Appendix S1.

We simulate using a single representative distribution of family sizes. We have checked that alternative realizations based on draws from the same distribution do not give qualitatively different results. Our choice of the workplace-size of the outbreak location is representative of a mid-sized farm or wet-market. This helps eliminate a major source of stochasticity – the location is large enough that we need not account for stochasticity in infections in the bird population, thus permitting the modelling of infections in humans acquired from birds using a deterministic force of infection.

### 2.3 Disease progression

The infection of primary contacts, humans in direct contact with infected birds, is described through a force of infection. This is depicted in Fig. 1(B) (the smooth curve). This force of infection in the outbreak site is taken to be proportional to the fraction of infected birds in that location. In our model of a farm-outbreak, we choose parameters of transmission among birds such that the force of infection arising from the birds peaks at approximately 12 days from the start of our simulations. The first primary case is detected close to the peak of the force of infection, about 10 days after the initiation of the infection in this run. In practice, information would be available, at least in principle, about the fraction of birds which die (containing information about to the force of infection and how it changes in time, the curve) and the time-line of cases in humans (circles and crosses).

Fig. 2 shows the disease progression in our model for H5N1. Disease states are described using the notation of conventional compartmental models. Given their high density, infection among birds is assumed to follow a variant of the well-mixed continuous SIR model, which we will from now on denote as the “SID” model. Susceptible birds (*S*_*B*_) can become infected (*I*_*B*_) before dying from the disease and transitioning to the *D*_*B*_ compartment. More details of this model can be found in Appendix S2.

**Fig. 2:**
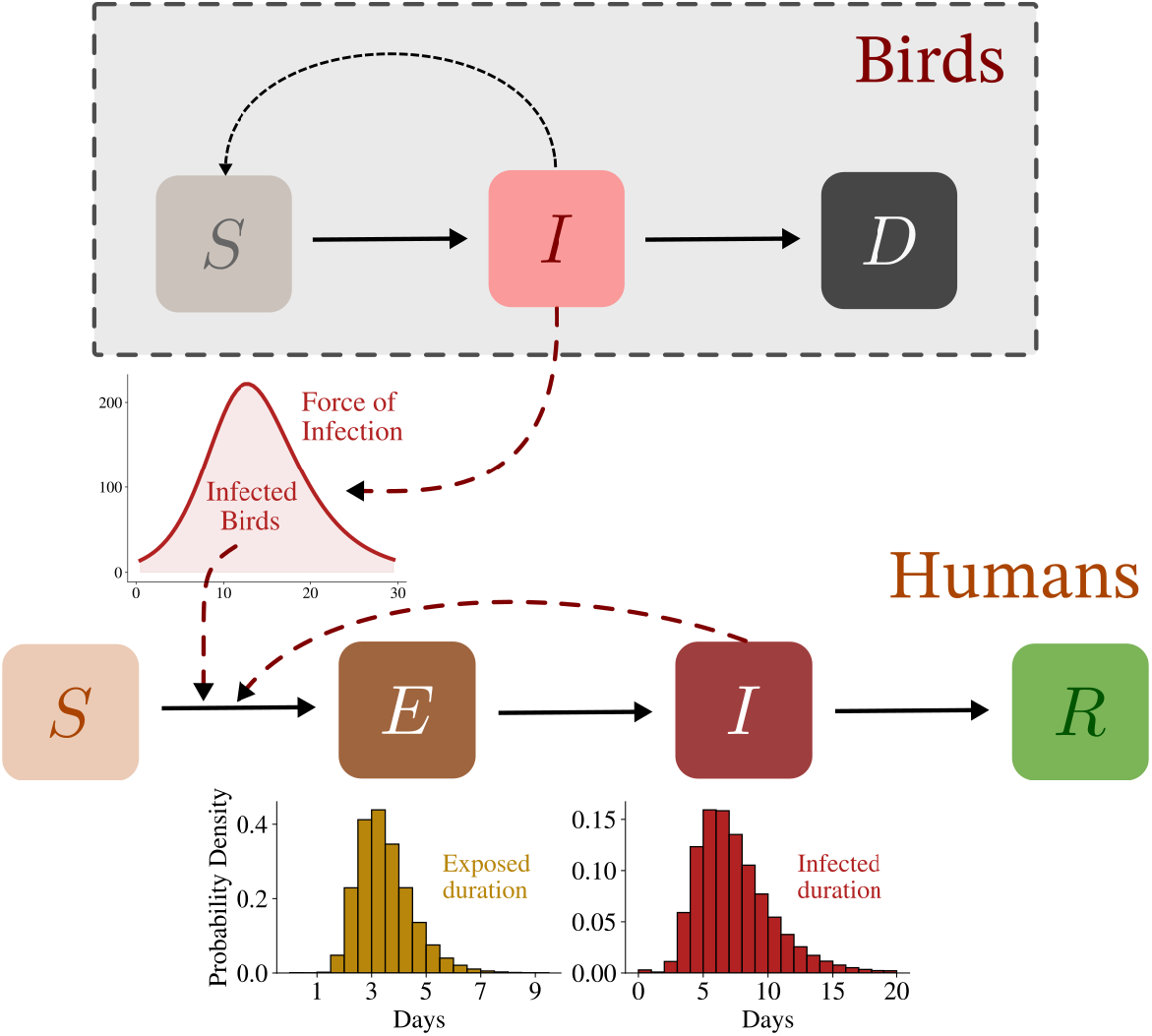
Description of the disease model. The birds are assumed to follow a well-mixed compartmental variant of the SIR model where they are infected and eventually die (which we denote as an “SID” model since the birds can be (S)usceptible, (I)nfected or (D)ead). The total number of birds contributes to the force of infection at the outbreak site. Individual agents can be in any one of the four states (S)usceptible, (E)xposed, (I)nfected, or (R)emoved. An agent working at the outbreak site may be infected by these birds, after which they transition to the “Exposed” state, and eventually become infectious and can infect their secondary (household) contacts who can, in turn, infect other contacts in the population.

For humans, even though they are described as individual agents, we use the *notation* of an SEIR model to classify their disease states. At any given time, each individual can be in one of four possible states: (S)usceptible, (E)xposed, (I)nfectious, and (R)emoved. The disease states are shown in Fig. 2. The disease is assumed to be symptomatic. We assume that recovered individuals are immune to further infection. Our model can be generalised to include the possibility of asymptomatic transmission. Human agents transition stochastically between disease compartments after a probabilistically drawn duration (or “sojourn time”). BharatSim allows modellers the flexibility of choosing these sojourn times from different probability distributions. We draw these times from lognormal distributions, whose properties are specified further in Section 2.4.

### 2.4 Parameters

Our model community consists of 9,667 individuals, constructed to simulate a catchment area for the given outbreak site. (Since our interest in the trajectory of initial infections, ranging between 0 and 100 typically, this choice is sufficient.) Agents employed by this site are primary contacts. The remaining network of individuals represents (i) their household members (in addition to other, secondary, contacts), and (ii) work- or school-contacts of their household members (tertiary contacts). Our parameter choices are detailed in Table 1.

**Table 1:** Parameter choices for our model. Our simulations are run using multiple parameter sets. Those used in this paper correspond to the parameter choices above.

| Par. | Description | Range of values | Comment |
| --- | --- | --- | --- |
| $\beta_{\text{BH}}$ | Bird-human transmission probability | $(1, 5, 10) \times 10^{-3}$ | Correspond to $\sim 20$, $\sim 70$, and $\sim 90\%$ probabilities of at least one spillover event. |
| $\beta_{\text{BB}}$ | Bird-bird transmission probability | 0.667 | Model choice, based on Ref. [16]. |
| $R_0^B$ | $R_0$ for bird SID model | 2.3 | Corresponds to a peak at $\sim 15$ days after initialisation; agrees with Refs. [16, 34]. |
| $N_{\text{pop}}$ | Human population size | 9,667 | Primary: 165, secondary: 476. |
| $\tau_E$ | Latent period for human infection | lognormal(3,1) | Drawn from mean of 3 days [12, 24] and standard deviation of 1 day (model choice). |
| $\tau_I$ | Infectious period for human infection | lognormal(7,1) | Drawn from mean of 7 days [12, 24] and standard deviation of 1 day (model choice). |
| $\beta_{\text{HH}}$ | Human-human transmission parameter | (0.05, 0.5) | Correspond to $R_0 \in (0, 3)$. |
| CD | Culling day | (10, 20, 600) | The FOI due to birds is set to zero post $t = \text{CD}$. |
| QT | Quarantine threshold | 2, 10 | Threshold number of infectious agents for lockdown of primary contact households. |
| DVR | Daily vaccination rate | 0%, 1%, 5% of $N_{\text{pop}}$ | Number of vaccines available per day. |
| VE | Vaccine efficacy | 0%, 50%, 100% | Reduces relative risk of infection by VE. |

For *β*_HH_, we choose values in a range that corresponds to a basic reproductive ratio *R*_0_ *∈* (0, 3). (This must be back-calculated given the trajectory of infections for a given *β*_HH_, as we detail below.) The distributions of sojourn times in each compartment (shown in Fig. 2) are uncertain at the outbreak onset. We draw them from lognormal distributions, choosing parameter values that should be largely representative [12]. In itself, this choice of distribution provides a considerable improvement on conventional compartmental models, where the sojourn times in each compartment are constrained to be exponentially distributed. These parameter choices will have to be refined as more information about the outbreak becomes available. Our simulation framework can easily account for updated parameter values.

### 2.5 Initialisation

We begin our simulations with zero infected individuals in the human population, but with a small number of infected birds in the outbreak site. Susceptible birds get infected at some rate *β*_BB_, and eventually die. At every time-step, individual agents who work in the farm are assumed to come in contact with these infected birds. The fraction of infected birds thus contributes to the force of infection within the outbreak site.

For human-to-human transmission to occur at all, a pre-requisite is an initial spillover event. To account for this, we weight the infected fraction of birds by a “relative risk of spillover”, *β*_BH_, which we vary in the range *β*_BH_ *∈* (10^−6^, 10^−5^). This corresponds to between 20% and 90% of the simulation runs leading to at least one spillover event, as we show in Appendix S3. Once such an event occurs, the disease is assumed to spread amongst the human population through direct contact. At any given time, each location within our simulation (home, work, school) is considered to be a fully-connected network of individuals. However, the number of individuals an agent comes in contact with changes with time as they move between these locations.

### 2.6 Force of infection

Susceptible individuals in each location experience a force of infection given by

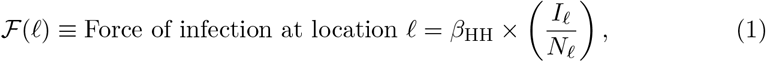

where *β*_HH_ is the human-human transmission parameter, *N*_*𝓁*_ is the total number of individuals in that location, and *I*_*𝓁*_ is the number of infected individuals in that location. In an interval of time Δ*t*, “Susceptible” individuals transition to the “Exposed” compartment with probability ℱΔ*t*.

We further consider the case of a constant force of infection at the work location of primary contacts. This reproduces a scenario where primary contacts have a risk of acquiring an infection which remains unchanged over an extended period of time. This would indicate exposure to a more-or-less constant number of infected birds. Such a scenario might be expected to be applicable to a wet market where birds and move in and out at constant rate, with a fixed fraction of them being infected.

To compare our results to those in the case of the farm-based outbreak model, the constant force of infection is set by ensuring that it has the same integral as the force of infection derived from the fraction of infected birds. Thus, we consider a constant force of infection given by

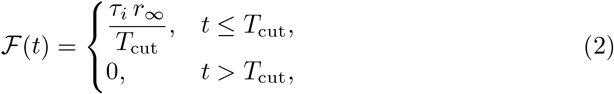

where *τ*_*i*_ is the mean infected duration of the birds, *r*_∞_ is the total outbreak size (or total number of infected birds) in the case of no culling as a fraction of the total bird population, and *T*_cut_ is a cutoff beyond which the force of infection drops to zero. We choose this cutoff to be 40 days, corresponding to the day when the integrated probability of exposure of a single primary contact is 99.7% of what it would have been in the no-culling farm-outbreak scenario.

### 2.7 Interventions

We consider three specific interventions and study their impacts on the disease trajectory.

#### Culling

At a specific day, all the birds in the farm are culled. Beyond this time, no further infections can arise from bird-human interactions. Any new cases after this date occur solely due to human-human interactions. This is modelled in our simulations by setting the force of infection arising from birds to zero, as shown in panels (C) and (E) of Fig. 3.

**Fig. 3:**
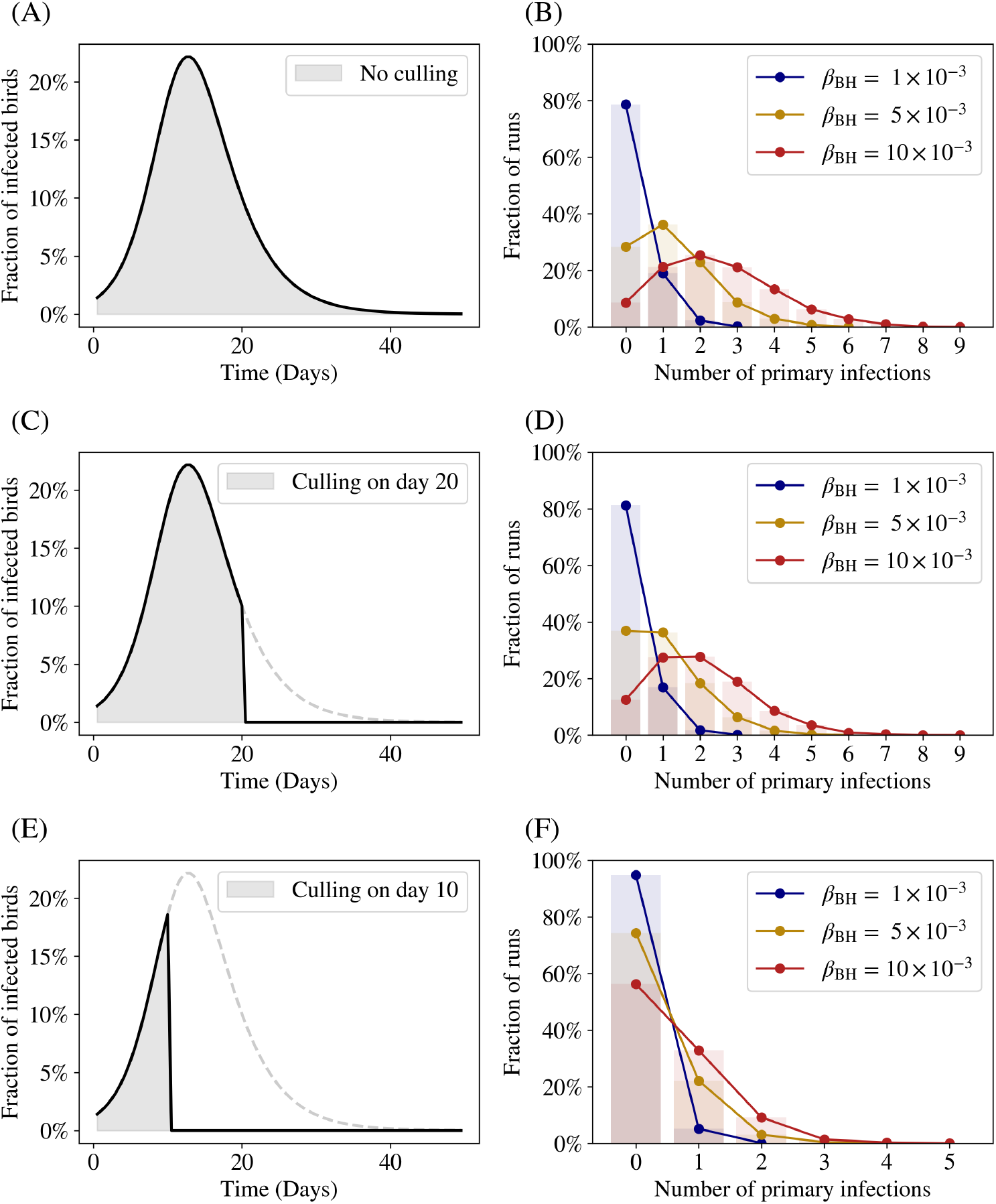
The distribution of primary cases. Each row represents a different culling date, with successive panels representing earlier culling dates, as shown in the panels (A), (C), and (E). The solid curve represents the number of infected birds in the farm, which abruptly goes to zero in the case of culling. The dashed grey curve represents the number of infected birds in the absence of culling. The right panels (B), (D), and (F), show the probability distributions of the number of primary cases for these culling dates, with the different colours representing different values of *β*_BH_, the bird-human interaction probability.

#### Vaccination

We consider a vaccination drive that begins one week after the detection of the first case in the population. Vaccines are restricted to primary and secondary contacts, and have the effect of reducing the agents’ susceptibility to the disease. Given that little is known about how such vaccines might function, we choose to assume that vaccination does not alter the infectivity of an individual who contracts the disease, vaccinated or not.

We consider a daily vaccination rate (DVR) of 1% of the population (which translates to 100 vaccines available for use every day). We choose vaccine efficacies of 50% and 100%, which corresponds to the reduction in susceptibility of a vaccinated agent. (A vaccine efficacy of 100% means that an individual is completely immune to the disease.) The effect of the vaccine is immediate.

We have also performed simulation runs for a DVR of 5%, and with a vaccine drive that begins two weeks after the first detected cases. These results are shown in Appendix S4.

#### Quarantine

When the number of cases reaches a threshold value (which we choose as either 2 or 10 individuals in our simulations, to explore a range of possible scenarios), all households of primary contacts are quarantined, such that no individual is allowed to leave their household to their workplaces or schools.

### 2.8 Computing *R*_**0**_ and the secondary attack risk

From our simulations, we compute two specific quantities: the basic reproductive ratio *R*_0_, the secondary (or household) attack risk (SAR). We also study the tertiary attack risk (TAR), described in detail in Appendix S4.

#### 2.8.1 Reproductive ratio *R*_**0**_

The basic reproductive ratio is defined as the average number of people infected by a single infectious person in the background of a population of completely susceptible people. Its value, in an averaged sense, is dependent on the mean number of contacts per unit time, the probability that an infection will result from an encounter with an infected contact over the period of the interaction, and the time over which an individual remains infected. Stochasticity enters through the network structure of interactions and the non-deterministic nature of the transmission of infection.

Our agent-based simulations take both of these into account when determining *R*_0_. The advantage of agent-based models is that these different sources of stochasticity can be tuned independently of each other, in contrast to purely statistical models.

To compute *R*_0_ from our simulations, we record all chains of infection. Whenever an individual is infected in a specific location, an agent is drawn at random from the potential “infectors” at that location. For a given value of *β*_HH_, we run our simulations for a period of 21 days from the date of spillover. This period is chosen so that a sufficient number of infectious individuals recover, while the number of susceptibles in the accessible population can be assumed to be more or less constant. At the end of this period, we compute the average number of people infected by a person who has recovered from the disease.

#### 2.8.2 Secondary or household attack risk (SAR)

The secondary attack risk measures the difference between community transmission of illness versus transmission of illness in a household and is a useful indicator of infectivity at outbreak onset. It is calculated as the proportion of infected household or family contacts [35]:

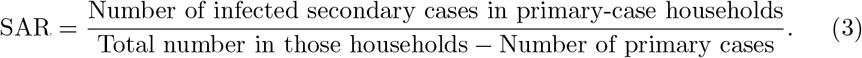

Consider an outbreak in which 5 “primary” cases are detected. After one incubation period, 10 new cases develop in the households of these primary contacts. If the combined total number of individuals in these households was 30, then the secondary attack risk is calculated as

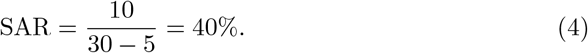

As with our computation of *R*_0_, we restrict our computation of the secondary attack risk to cases within the first 21 days of the first recorded case. In this time, it is unlikely that *all* households containing primary contacts have an infection. As a result, the secondary attack risk uses only those households in which one primary case has been detected. Additionally, in our computation of the SAR, we do not differentiate between infections in primary contacts that arise due to interaction with the FOI, and those that are due to interactions with other infected primary contacts.

#### 2.8.3 Tertiary attack risk (TAR)

We also compute the tertiary attack risk (TAR), which measures the likelihood of transmission to third-generation cases, i.e., those individuals infected by secondary cases. The TAR describes the potential for sustained transmission beyond initial household exposure, providing insight into transmission dynamics outside the immediate sphere of influence of primary-contacts. The TAR is particularly useful in evaluating the effectiveness of interventions and the potential for outbreak amplification, as it extends the scope of potential infections to the workplaces of secondary contacts.

Following analogous definitions in household transmission studies [35], the TAR is calculated as:

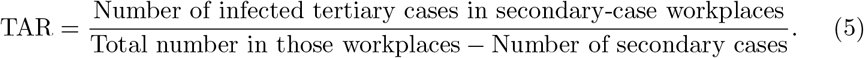

## 3 Results

We begin our simulation with no infected humans and a small fraction of infected birds. The infection spreads rapidly amongst the birds and potentially infecting the farm workers (the primary contacts). In this section we study the effect of different bird-human interaction probabilities, culling days, and quarantine strategies on the disease trajectory.

### 3.1 Distribution of primary cases

We begin by studying the distribution of primary human spillover cases in different scenarios of bird-culling. We focus on estimating the number of infected primary contacts as a function of two specific parameters: the day on which the birds are culled, and the bird-human interaction parameter *β*_BH_.

We consider the case when the force of infection on the primary contacts is proportional to the fraction of infected birds, scaled by the factor *β*_BH_. In principle, we could account for changes in the force of infection due to changes in human behaviour, such as the use of personal protective equipment (PPEs), but we do not do so here. In panels (A), (C) and (E) of Fig. 3, we show this fraction arising from the spread of disease in the bird population. In the absence of culling, the force of infection follows the dashed line. The solid line shows the fraction of infected birds when all birds are culled on three specific days following the onset of the infection.

In panels (B), (D), and (F) of Fig. 3, we show the distribution of primary cases for three values of the human-bird infectivity. The different panels show the results for different culling scenarios. In each case, we run 500 stochastic simulations on the same population to generate the distributions of the primary cases. We find that for the earliest culling dates, the distribution of primary cases decreases monotonically. The most likely outcome is that there are no primary cases, with a very low probability of one or more than one spillover event.

With delayed culling, we find that the prolonged exposure of the primary contacts with the bird-population shifts the distribution of primary cases rightwards. Thus, for later culling dates, the distribution of primary cases peaks at 1 or 2 individuals, but also has a long tail, where as many as 7 or 8 individuals may be infected. As *β*_BH_ increases, the peak of this distribution shifts outward, while its spread becomes larger.

The results in Fig. 3 highlight the amplification of spillover risk as transmission from birds persists, and highlights the importance of avian control measures in curtailing the risk of a spillover event.

In the case of a uniform force of infection, argued to be relevant to the case of a wet-market, we find that the distribution of primary cases matches very closely with the farm-outbreak scenario for both the early-culling and no-culling scenarios. For other scenario, at intermediate culling times (i.e. for culling dates between 10 and 30 days) and for low values of *β*_BH_, both scenarios continue to match. However, the distributions of primary cases in each of these cases show some differences when *β*_BH_ becomes larger, at these intermediate culling times. A more detailed analysis of the primary-case distribution for intermediate culling-dates, including a theoretical computation for this distribution, can be found in Appendix S5.

The results of Fig. 3 illustrate both the impact of increasing the probability of a spillover event as well as the effect of culling on the distribution of primary human cases. We find that culling has a large effect on this distribution, provided it occurs sufficiently early, before the peak in the force of infection arising from the infected birds. In this case, the most likely scenario is that the infection will not spill over, irrespective of bird-human spillover *β*_BH_. For later culling dates, the most likely scenario involves one or more primary cases, a number that depends strongly on *β*_BH_. This argues for early culling as an important strategy to curb further spillover events.

### 3.2 Reproductive ratio and secondary attack risk

Having established the potential for potential zoonotic spillover events under the different conditions described above, we next examine the potential for sustained human-to-human transmission by estimating two epidemiological metrics, the basic reproductive ratio (*R*_0_) and the secondary attack risk SAR, computed from the results of our simulations.

In an agent-based simulation, the basic reproductive ratio is not simply specified in terms of the model parameters, because it depends on the assumed network structure. To account for this, we compute *R*_0_ directly from the distribution of secondary infections. This follows the exact definition of *R*_0_ in terms of the average number of secondary cases caused by one infected individual in an otherwise susceptible population. While other, purely statistical, methodologies for calculating *R*_0_ from a time-series of cases could be applied here, our calculation is sensitive to the contact network structure of our population, a relevant detail that generic statistical methods cannot capture.

Figs. 4 and 5 show *R*_0_ for different values of the human-human transmission coefficient (*β*_HH_), where the error-bars arise from the inherent stochasticity of the transmission process: these graphs are obtained by averaging over multiple stochastic runs. In principle, one might have to average over different network structures as well, but here we assume that a well-constructed synthetic population should naturally improve on the conventional approximation of a well-mixed population generally used in traditional compartmental models.

**Fig. 4:**
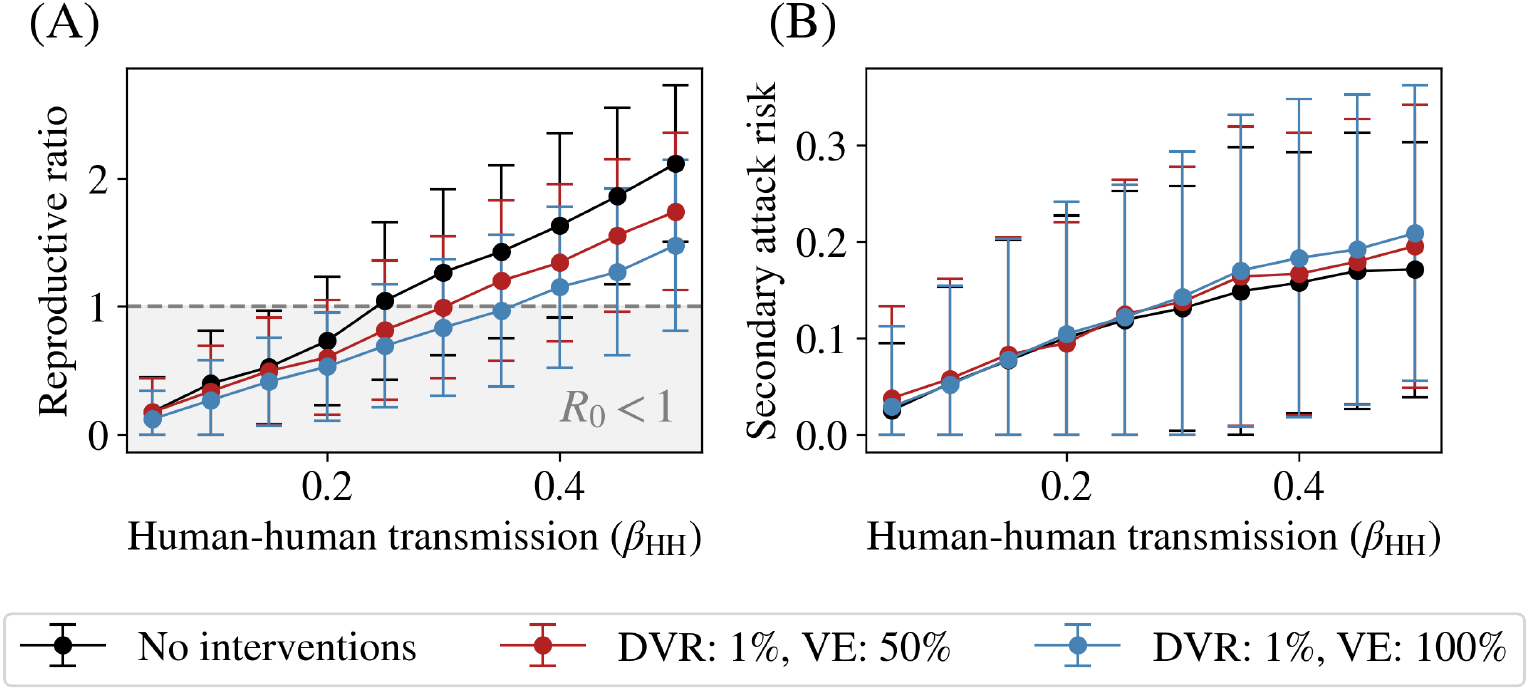
Calibrating the human-human transmission rate. (A) The mean repro-ductive ratio as a function of the human-human transmission *β*_HH_. We see that in our model, *β*_HH_ ≈ 0.25 represents the threshold *R*_0_ = 1, beyond which the epidemic takes off among humans in the case of no interventions. The introduction of vaccinations causes the critical threshold of *R*_0_ = 1 to shift rightwards towards higher values of *β*_HH_. (B) The secondary attack risk (SAR) among secondary contacts, evaluated as described in Equation (3). We see that the introduction of vaccinations does not alter the secondary attack risk. The error bars in both plots represent the variation (one standard deviation) across 500 different stochastic runs.

**Fig. 5:**
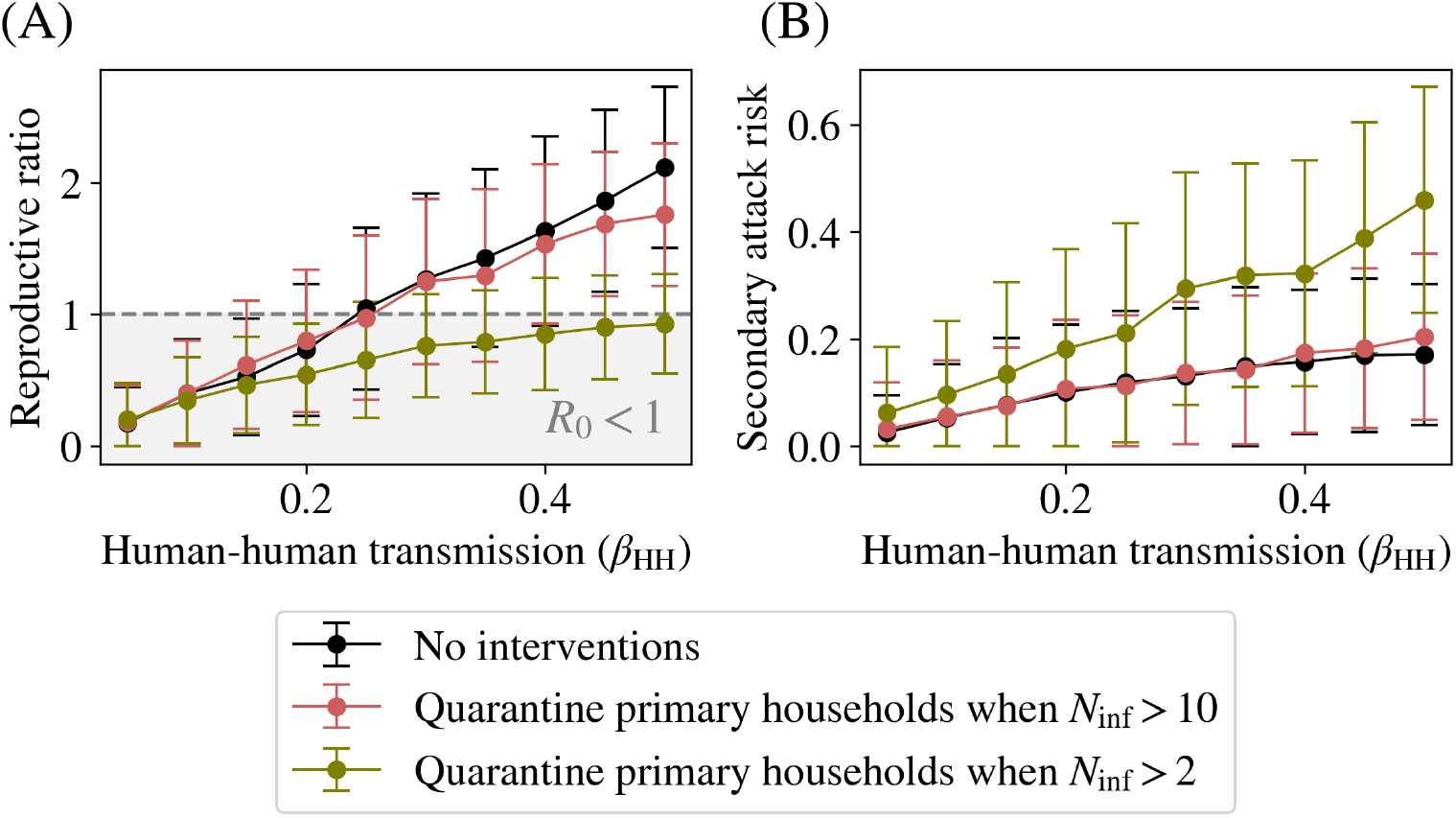
Effects of quarantining strategies. (A) The mean reproductive ratio as a function of the human-human transmission *β*_HH_ in the case of quarantining homes of farmers when a threshold of infections is reached. As in Fig. 4, *β*_HH_ ≈ 0.25 represents the threshold *R*_0_ = 1 in the case of no interventions. However, if farmers and their households are quarantined when either 2 or 10 infected cases are detected, this can lead to a reduction in the number of cases within primary and secondary contacts. (B) The secondary attack risk (SAR) among household contacts of farmers. The quaran-tining of farmer households greatly reduces the SAR, although it does not necessarily reduce the spread of infection in the population unless it is done early. As can be seen, using a threshold number of cases of 2, for example, leads to an epidemic with an *R*_0_ consistently below 1, meaning the epidemic never takes off. The error bars in both plots represent variation (one standard deviation) across 500 different stochastic runs.

In Figs. 4(B) and 5(B), we show the secondary attack risk as a function of *β*_HH_, and show how the SAR may be used to infer *R*_0_. Given an estimated value of the SAR, we can compute the corresponding value of *β*_HH_ and hence the basic reproductive ratio *R*_0_ by simply reading off the curve. Both quantities are monotonic in the human-human transmission parameter *β*_HH_. While the structure of these curves depends in detail on the assumptions we make, such as for the parameters of the lognormal dis-tribution assumed for sojourn times in disease states, further refinements are possible as more clinical information is available to be assimilated during the early course of the outbreak.

In Appendix S6 we show the epidemic curves with a variety of interventions for five different values of *β*_HH_ that correspond to an *R*_0_ *>* 1. In addition to the no-intervention scenario, we also show the epidemic curves for a daily vaccination rate of 1% and a vaccine efficacy of 100%, and quarantining scenarios where the threshold is 10 and 2 individuals respectively.

Given the wide range of disease parameters in humans due to the paucity of avail-able data, we also performed a sensitivity analysis by choosing values of the serial interval in humans that were (i) larger by 25% and (ii) smaller by 25%. We repeated the runs described in the paper, and found no qualitative differences. The results of this analysis are shown in Appendix S7.

In Figs. 4 and 5, we show how the secondary attack risk can be used to infer the basic reproductive ratio, a central quantity in determining the pandemic potential of our outbreak. The reproductive ratio rises smoothly with *β*_HH_. In Fig. 4, we show the results with a vaccination drive. Introducing vaccinations causes the critical threshold of *R*_0_ = 1 to shift towards higher *β*_HH_, by an amount determined by the vaccine efficacy. We further find that the secondary attack risk is left relatively unchanged by the vaccination drive. This is due in part to our assumption that vaccination does not reduce the infectivity of an individual, only their relative risk of infection.

In Fig. 5, we show a similar graph but for quarantining primary and secondary contacts at different threshold values. As described above, when the number of active cases in the population exceeds a threshold of either 2 or 10 cases, the households of all primary contacts are locked down, and these agents are not allowed to travel to their workplace or school locations.

We find that a threshold of 10 infected individuals produces both an *R*_0_ and an SAR that is comparable to the case of no interventions. We attribute this to the fact that the time taken for 10 active cases to be present in the population is much larger than the 21 day interval used to compute the SAR, which is typically a quantity calculated early in an outbreak. In the case of a threshold of 2 infected individuals, we see a larger 16 SAR. This is because households of primary contacts are quarantined early on, and our models do not account for any reduction in spread within households. Consequently, infected primary contacts have a much higher probability of transmitting the disease to the secondary contacts in the households as they spend more time within the house. Nevertheless, the reproductive ratio remains consistently below *R*_0_ = 1, meaning that the epidemic does not take off, since the disease is not allowed to enter the tertiary contact network due to the quarantine. This is verified in Appendix S4, where we compute the tertiary attack risk explicitly.

Thus, we find that while quarantining households of primary contacts has by far the largest effect in reducing the spread of the disease, this must be done very soon after the first case is detected, failing which the disease will enter the tertiary contact network and a more aggressive lockdown strategy would be required. The error bars in both plots represent variation across 500 different stochastic runs.

We have also conducted runs combining these different interventions, as shown in Appendix S4, to show how the combined effects of vaccinations and quarantining primary and secondary contacts can affect these epidemiological parameters.

## 4 Discussion

In this paper we showed how computational models can describe the sequential stages of a zoonotic spillover. We showed that a recently developed, ultra-large-scale agent-based simulation framework for infectious diseases in an LMIC context, *BharatSim*, could be used to study the impact of various policy interventions in the case of an epidemic of a spillover event of HPAI. We modelled the possibility of initial spillover events of H5N1 from birds to humans, followed by sustained human-to-human transmission. Our model describes the two-step nature of outbreak initiation, showing how crucial epidemiological parameters governing transmission can be calibrated given data for the distribution of the number of primary and secondary cases at early times.

We considered the initial stages of an outbreak, reasoning that confining ourselves to simulating close to 10,000 individuals overall should be sufficient. The world’s fastest growing poultry markets are in South-and South-East Asia [36, 37]. One feature of poultry farms in these regions is a coexistence of small family-holdings with large-scale factory-farmed poultry. Since large factory farms are extensively monitored, the more interesting situation for us is the intermediate-sized one. We ignore stochastic effects in the bird population in the limit we consider, thus limiting sources of randomness.

The range of estimated epidemiological disease parameters in humans is very broad, given the sparsity of data. We used the recent meta analysis of Ref. [12] to fix values for the incubation and infectious periods. Changes in these within the estimated ranges will not affect our qualitative results.

Our simulations account for different interventions. These include (i) the culling of all birds in the farm, (ii) the quarantining of primary and secondary contacts once a threshold of cases is crossed, and (iii) a vaccination drive where primary and secondary contacts are targeted. We find that culling birds is effective, provided no primary infection has occurred. The earlier birds are culled, the larger the probability that a spillover can be prevented. We further compare two scenarios: in the first, simulating a farm-outbreak, the force of infection follows an SID model with a characteristic peak where the number of infected birds is largest. A second scenario considers the case of the wet-market. Here, as we argued, we model the balance of the birds entering and leaving as leading to a flat force of infection. The total area of infection curves in both cases is normalised to be the same, and represents the total number of bird-hours of contact with infected birds. In both cases, culling early reduces the chances of a spillover event. The detailed structure of the force of infection matters most at intermediate culling dates where the areas of the FOIs prior to culling differ maximally. In our study of the tertiary attack risk, we found that even if an infection of a primary case occurs, onward infections are limited if cases are isolated and their household contacts quarantined. However, once tertiary contacts are infected, establishing control becomes impossible unless far more stringent measures are applied, including a total lockdown.

As expected, the reproductive ratio in humans rises smoothly with the human-to-human transmission parameter *β*_HH_. Introducing a targeted vaccination drive aimed at primary and secondary contacts causes the critical threshold of *R*_0_ = 1 to shift towards higher *β*_HH_, by an amount determined by the vaccine efficacy. The secondary attack risk is left relatively unchanged by the vaccination drive. This is due in part to our assumption that vaccination does not reduce the infectivity of an individual, only their relative risk of infection.

We also considered the effect of quarantining households of primary contacts on the secondary and tertiary attack risks. We find that a quarantine threshold of 10 infected individuals produces both an *R*_0_ and an SAR that is comparable to the case of no interventions. We attribute this to the fact that the time taken for 10 active cases to be present in the population is much larger than the 21 day interval used to compute the SAR, which is typically a quantity calculated early in an outbreak. The figure of 10 depends in detail on our choice of parameters for the serial interval of the disease in humans. A shorter serial interval should reduce this threshold. Infected primary contacts have a much higher probability of transmitting the disease to the secondary contacts in the households as they spend more time within the house. Nevertheless, the reproductive ratio remains consistently below *R*_0_ = 1, meaning that the epidemic does not take off, since the disease is not allowed to enter the tertiary contact network due to the quarantine. We have also investigated the synergies between different interventions, as discussed in Appendix S4.

The limitations of our methods are the following: our agent-based model assumes a specific, known, distribution of family sizes, fixes the sizes of workplaces and schools, and models the nature of contacts within them. These are determined by the detailed structure of the synthetic population. We also assume a dynamics in which agents move between their homes and workplaces or schools every 12 hours. These assumptions may not generalise across diverse geographical contexts, restricting the applicability of our results. Migratory birds and poultry distribution networks could potentially seed the infection at multiple nearby locations [38], a possibility we did not account for here. Although we know that a large fraction of poultry holdings are small, we chose to model an intermediate-sized farm so that we could ignore the stochasticity of infections in birds. Additionally, in our analysis, we ignore behavioural changes in primary contacts after the epidemic in birds is detected (such as the use of PPEs), although this is in principle easy to incorporate into our model.

It is in the very early stages of an outbreak that control measures make the most difference. Once community transmission takes over, cruder public-health measures such as lockdowns, compulsory masking, and large-scale vaccination drives are the only options left. At this stage, the large number of cases ensures that stochastic effects should play a smaller role and conventional compartmental models should provide appropriate guidance. However, projecting the trajectory of a disease from initial fragmentary information available at outbreak onset is a far harder task. It in this setting that the methods described here are expected to be most useful [31].

Conventional models that calculate the reproductive ratio from case data attempt to reproduce the observed trajectory of infections using statistical assumptions [39, 40]. This assumption collapses many independent sources of stochasticity, including network effects and the random nature of the transmission of infection. An agent-based model allows one to separate these different contributions. Because many parallel scenarios can be explored in real-time, and the models can be adapted to the measures that are actually implemented, such methods provide powerful input to policy [41]. Our methods allow us to explore a range of possibly applicable policy measures.

Understanding the scope of possible interventions in advance of a potential outbreak strengthens the ability of a public health infrastructure to respond. We know of no other work which has attempted to capture the consequences of such a two-step spillover event occurring in a realistic population modelled at the individual level, especially in the LMIC context. Our simulations can be run in real time, responding to initial reports of cases. They can be tuned further as more information is collected. Further generalizations include the incorporation of asymptomatic infections and of delays in reporting, the possibility of a bird to intermediate mammal to human chain of spillovers, the inclusion of serological data in real time, and a more refined description of the consequences of vaccination drives, including model descriptions of vaccine efficacy. These are the subject of our ongoing work.

## Data Availability

The simulation codes used to generate all figures are available on Github. The simulation framework is available at bharatsim.ashoka.edu.in

## 5 Declarations

### 5.1 Ethics approval and consent to participate

Not applicable.

### 5.2 Consent for publication

Not applicable.

### 5.3 Availability of data and materials

The simulation data on which this paper is based is generated by a model written in the freely available open-source simulation framework *BharatSim* (bharatsim.ashoka.edu.in). All presented results can be reproduced from the model code and the synthetic population file used to generate this data. Both have been made available on the GitHub repository https://github.com/dpcherian/h5n1-spillover-model.

### 5.4 Competing interests

The authors declare no competing interests.

### 5.5 Funding

Not applicable.

### 5.6 Authors’ contributions

Both authors did the literature search, design and implementation of the study, and wrote the original draft. Both authors had final responsibility for the decision to submit for publication.

## 5.7 Acknowledgements

The authors are grateful for ongoing support from the Mphasis F1 Foundation. Bharat-Sim development was supported by the Bill and Melinda Gates Foundation, Grant No: R/BMG/PHY/GMN/20, as well as by the Mphasis F1 Foundation. GIM acknowledges additional support from the National Disease Modelling Consortium at the Indian Institute of Technology, Bombay. The authors acknowledge the use of computational facilities provided by the Centre for Bioinformatics and Computational Biology, as well as the One-Health initiative of the Center for Climate Change and Sustainability at Ashoka University. PC would like to acknowledge Bhavesh Neekhra for his help with the generation of the synthetic population. The funders had no role in the study design, data collection and analysis, decision to publish, or preparation of the manuscript.

## Supporting information

## Appendix S1

### Description of the synthetic population

We describe synthetic population used to generate the results in the main paper.

## Appendix S2

### Equations for the well-mixed SID model for birds

We describe the compartmental model that is used to model the spread of the disease within the bird population.

## Appendix S3

### Calibrating the bird-human interaction parameter

We show how we calibrate the bird-human interaction parameter *β*_BH_ using the spillover probability.

## Appendix S4

### Synergies of different intervention strategies

We compare the epidemiological parameters *R*_0_, SAR, and TAR for different combinations of interventions.

## Appendix S5

### Comparing different forces of infection

We compare the effect of different FOIs on the primary case distribution.

## Appendix S6

### Epidemic curves with different interventions

We compare the different epidemic curves for different values of *β*_BH_, *β*_HH_, and different intervention strategies: no interventions, a vaccine drive with a daily vaccination rate of 1%, and quarantining all primary and secondary contacts when the number of infected agents in the population is greater than 10 or 2 respectively.

## Appendix S7

### Sensitivity analysis for epidemiological parameters

We perform a sensitivity analysis by varying the incubation and infectious period of the disease in humans and comparing the results to those from the main paper.

## Notes

### Competing Interest Statement

The authors have declared no competing interest.

### Funding Statement

The authors are grateful for ongoing support from the Mphasis F1 Foundation. BharatSim development was supported by the Bill and Melinda Gates Foundation, Grant No: R/BMG/PHY/GMN/20, as well as by the Mphasis F1 Foundation.

### Summary of Updates

Expanded text including discussion section and supplementary information, added discussion regarding wet markets

