## Supplementary Information for "Modelling a potential zoonotic spillover event of H5N1 influenza"

### S1 Appendix: Description of the synthetic population

For our study, we restrict ourselves to a small village in the district of Namakkal in the state of Tamil Nadu in India. We use the BharatSim synthetic population pipeline to generate a population that accurately reflects the demographic structure and household composition of this district. Namakkal is a key poultry-producing region with a high density of backyard and commercial poultry farms, making it a plausible site for zoonotic spillover of avian influenza such as H5N1. By simulating transmission in a realistic, spatially granular population, our model can potentially capture heterogeneities in contact networks, household structure, and vaccine allocation that are critical for evaluating the risk of local amplification and the effectiveness of targeted interventions.

Aggregate data from the 2011 Census of India is used along with a statistical sampling technique called Iterative Proportional Updating (IPU) which accepts both information regarding marginals from the real population, and individual-level data from a sample of the real population. IPU is used to iteratively scale up the sample to generate a large-scale population, ensuring that the marginal distributions for both households and individuals in the synthetic population match with the known marginal distributions in the real population.

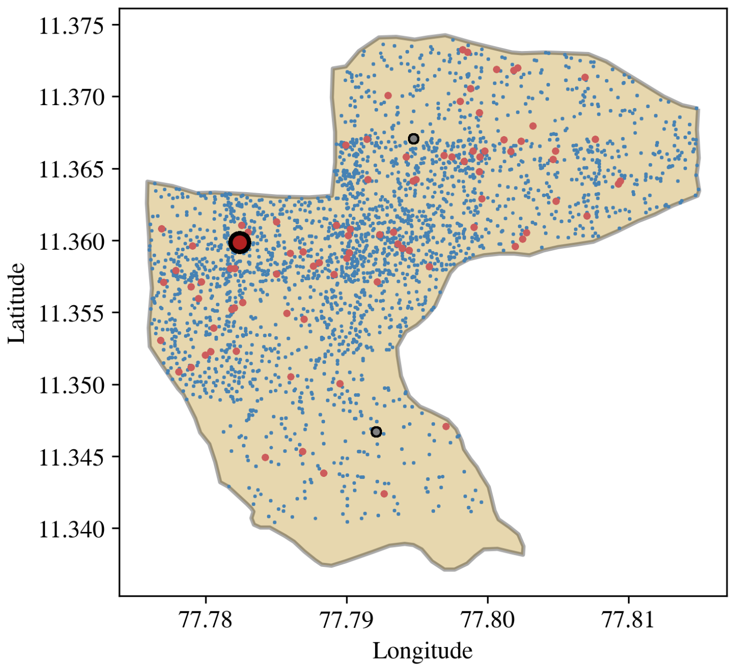


**Fig S1.1: Geographical distribution of population.** The geographical distribution of homes (small blue dots), workplaces (larger red dots), and schools (larger grey dots) are shown overlaid on the shapefile for Kadachanallur. The farm is denoted by the large red circle on the top left of the plot.

We use a subset of the IHDS-II dataset obtained by filtering for individuals and households which are situated in the state of Tamil Nadu. (We use the filtered datasets for the entire state, since the district dataset in IHDS-II contains very few samples, which are not enough to be able to generate a quality synthetic population.) This data is also used to draw a job label for every individual, satisfying the empirical distribution observed in the IHDS-II subset for Tamil Nadu.

We choose an arbitrary, small, village in the district of Namakkal that has a population of roughly 10,000 individuals in the 2011 Census data. In this study, the village we chose is the village of Kadachanallur. Using the shapefile for this district, we generate synthetic workplaces, homes, and schools and distribute them based on the data from the Gridded Population of the World. Individuals in our synthetic population are associated with external locations which include workplaces, schools, and public places. These locations define their contact networks. We designate a location containing agents with a job label of "Farm worker" to be a poultry farm. This is shown in Fig. S1.1.

All individuals are assigned job descriptions. This job description is sampled with replacement from the list provided in the IHDS-II dataset. Individuals below the age of 3 are "Homebound". For individuals above the age of 3 and below the age of 18, "Student" is assigned as the job description. Close to 30% of our population is below the age of 20, as can be seen in Fig. S1.2.

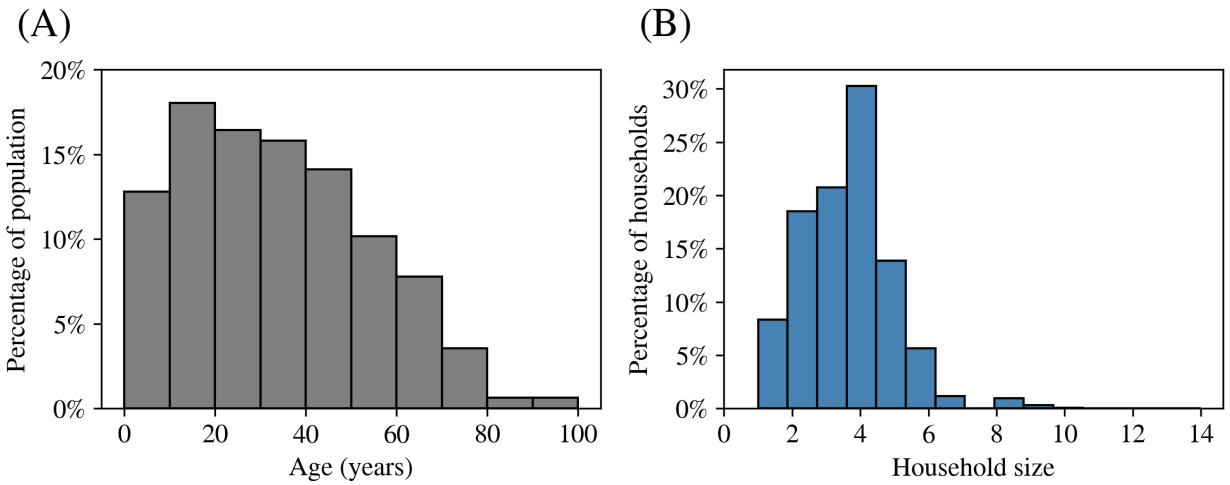


**Fig S1.2: Age-distribution and household-distribution in our synthetic population.** (A) shows the fraction of agents in each 10-year age-band, and (B) shows the distribution of household sizes.

One of the challenges in making a synthetic population is the need for up-to-date survey data. The most recent iteration of our main source of microdata, the IHDS-III survey, was expected to be released in 2023, but has still not yet made public. Additionally, the last India-wide census was conducted in 2011. The absence of up-to-date census data has made the task of extrapolating to the current date far more difficult. Following most, if not all such modelling efforts, we have used data from the 2011 census to generate the population used in this paper.

### S2 Appendix: Equations for the well-mixed SID model for birds

The SID model described in this paper is a compartmental model in epidemiology that divides the population into three mutually exclusive compartments:

- $S(t)$: the number of susceptible birds at time $t$,
- $I(t)$: the number of infectious birds at time $t$,
- $D(t)$: the number of deceased birds at time $t$.

We replace the standard "Recovered" compartment with a "Dead" $D(t)$ compartment, since HPAI has historically caused 75-100% mortality in birds. The model assumes a closed population and describes disease spread using the following set of ordinary differential equations (ODEs):

$$\frac{\mathrm{d}S}{\mathrm{d}t} = -\beta_{\mathrm{BB}}\left(\frac{SI}{N}\right),$$

$$\frac{\mathrm{d}I}{\mathrm{d}t} = \beta_{\mathrm{BB}}\left(\frac{SI}{N}\right) - \gamma_{\mathrm{BB}}I,$$

$$\frac{\mathrm{d}R}{\mathrm{d}t} = \gamma_{\mathrm{BB}}I,$$

where $\beta_{\mathrm{BB}}$ is the transmission rate, $\gamma_{\mathrm{BB}}$ is the recovery rate, and $N = S + I + R$ is the total population (assumed constant).

The basic reproduction number is defined as $R_0 = \beta_{\mathrm{BB}}/\gamma_{\mathrm{BB}}$, representing the expected number of secondary infections from a single infected individual in a fully susceptible population.

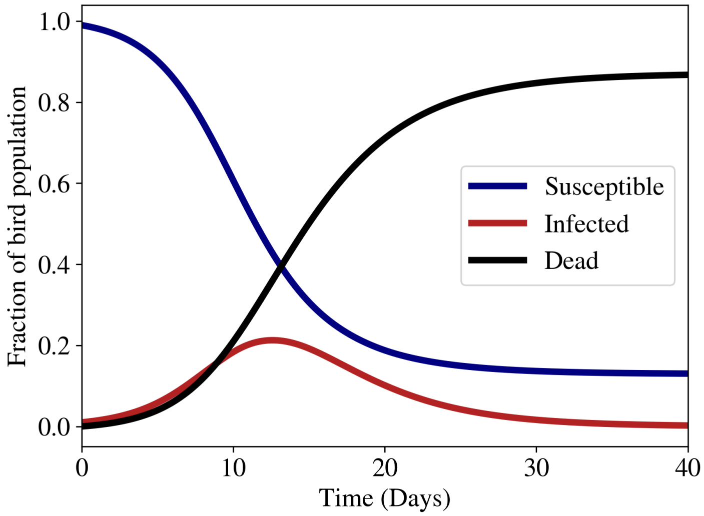


**Fig S2.1: Epidemic curves for the well-mixed bird population.** We consider a Susceptible-Infected-Dead model for the bird population.

### S3 Appendix: Calibrating the bird-human interaction parameter

As described in the main text, the bird-human interaction parameter range is chosen by computing the number of spillover events that each value could produce. The three values chosen in this paper, $1 \times 10^3, 5 \times 10^{-3}$, and $10 \times 10^{-3}$ correspond to roughly 20%, 70%, and 90% of runs leading to at least one spillover event respectively. This is shown in Fig. S3.1.

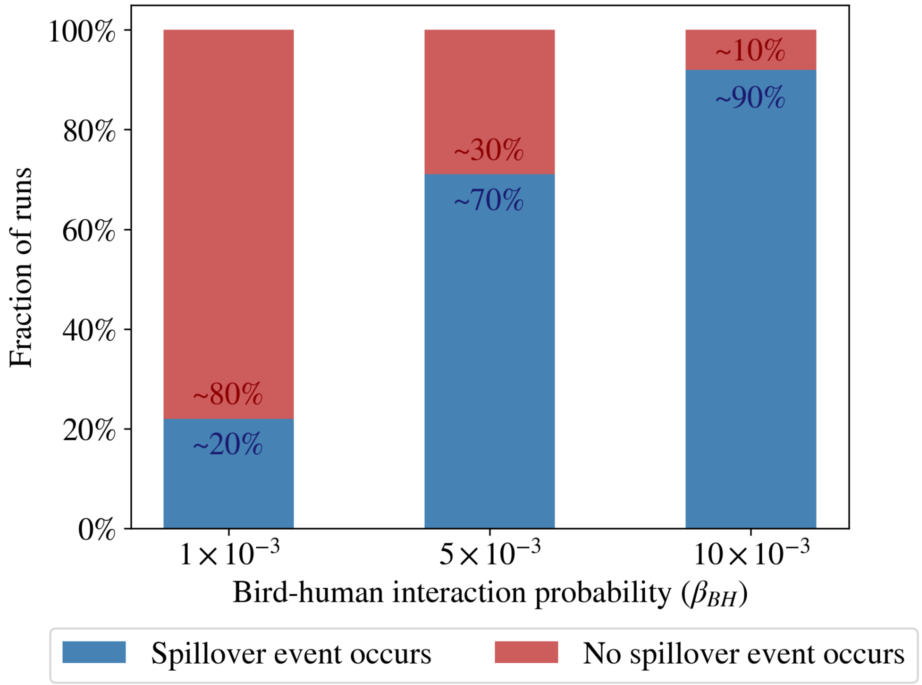


**Fig S3.1: Calibrating $\beta_{\mathrm{BH}}$ from probability of spillover.** The parameter $\beta_{\mathrm{BH}}$ controls the relative-risk of primary contacts from the FOI due to the infected fraction of birds in the farm. We choose three values of this parameter, to simulate runs which have a 20%, 70%, and 90% probability of spillover.

### S4 Appendix: Synergies of different intervention strategies

In this section we compare the reproductive ratio ($R_0$), the secondary attack risk (SAR), and the tertiary attack risk (TAR) for different interventions and their combinations. We compute the tertiary attack risk by computing the fraction of infected tertiary contacts, just as the secondary attack risk is the fraction of infected secondary contacts.

In Fig. S4.1, we show $R_0$, the SAR, and the TAR in the case of different quarantining strategies, i.e. when the threshold number of infected agents at which quarantine begins is 10 and 2 respectively. We see, just as in Fig. 5 of the main text that quarantining when the threshold is 10 reduces the number of secondary cases, but has little effect on the tertiary cases, since by that time the disease has escaped into the tertiary contact network, as is evidenced by $R_0$ continuing to be above 1. However, quarantining earlier keeps the reproductive ratio below the threshold of $R_0 = 1$, and allows for a TAR close to 0.

We also consider combinations of different interventions to explore synergistic effects. In Figs. S4.2 and S4.3 we see how combinations of the daily vaccination rate and quarantining strategies together affect the SAR and TAR. In these graphs, the vaccine drive is started one week after the first case is detected, meaning the vaccine delay is 7 days. In Figs. S4.4 and S4.5 we show the same graphs, but for the case of a vaccine delay of 14 days.

#### 4.1 No vaccination

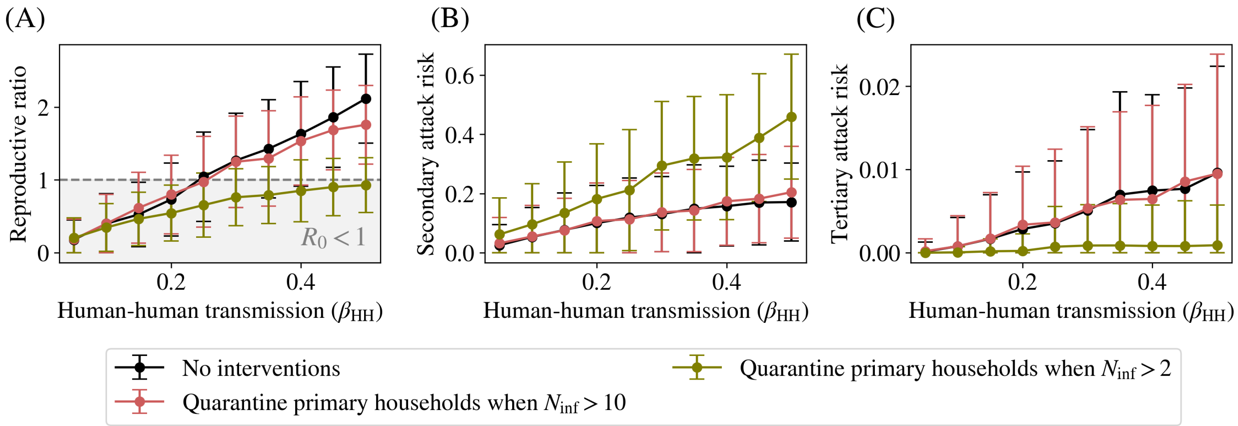


**Fig S4.1: Effects of quarantine on secondary and tertiary attack risk.** As shown in Fig. 5 of the main text, quarantine can reduce the risk of cases in primary and secondary contacts by bringing down the secondary attack risk. Here, we also show the same two panels (A) and (B) of this figure, but also add panel (C) which represents the *tertiary* attack risk amongst the rest of the population. The two quarantining strategies (when primary and secondary contacts are quarantined when 2 or 10 infectious individuals are detected in the population respectively) are both very efficient in curtailing the spread of disease amongst the primary and secondary contacts. However, its effect in reducing the TAR is highly dependent on when the quarantine is effected. While quarantining when 10 infectious individuals has almost no effect on the TAR, quarantining when 2 cases are detected stops the infection from spreading completely, keeping $R_0 < 1$. The error bars represent the variation (one standard deviation) across 500 different stochastic runs.

#### 4.2 Vaccine delay of 1 week

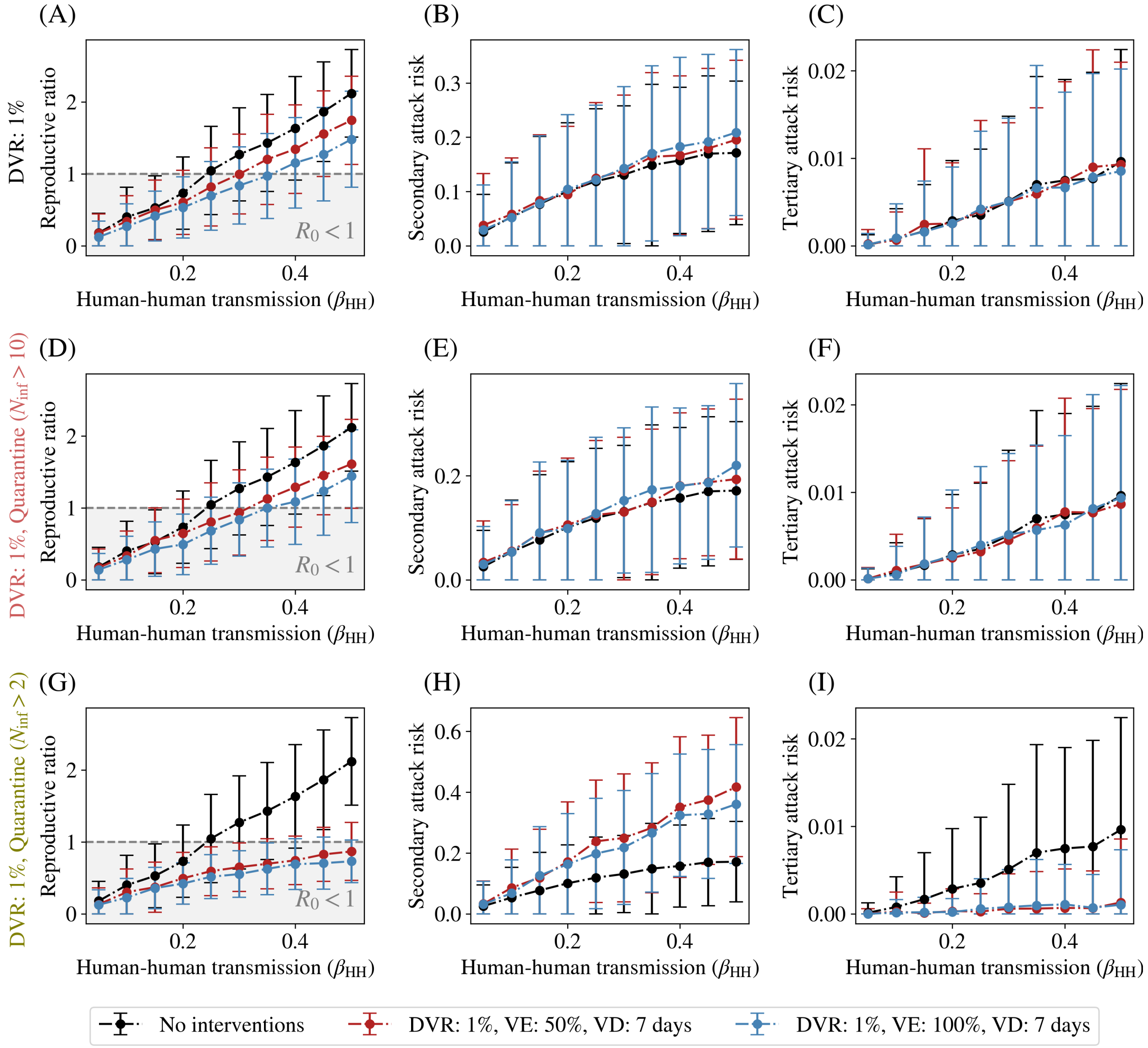


**Fig S4.2: Daily vaccination rate of 1% and delay of 1 week.** As shown in Fig. 4 of the main text, vaccination can reduce the risk of cases in primary and secondary contacts by bringing down the secondary attack risk. Here, we also show the same two panels (A) and (B) of this figure, and include panel (C) which represents the *tertiary* attack risk amongst the rest of the population. We can see that since vaccination is restricted to the primary and secondary contacts, and begins 7 days after the first infectious case has been detected, there is a high probability of the disease nevertheless spreading to the rest of the population, as evidenced by the high tertiary attack risk. While higher vaccine efficacy does bring this number down, the effect is far less substantial among the tertiary contacts when compared to the secondary contacts. In panels (D), (E), and (F) show the same curves when the primary and secondary contacts are quarantined when 10 infected agents are detected, while panels (G), (H), and (I) show similar graphs, but for a threshold number of 2 infected agents. The error bars represent the variation (one standard deviation) across 500 different stochastic runs.

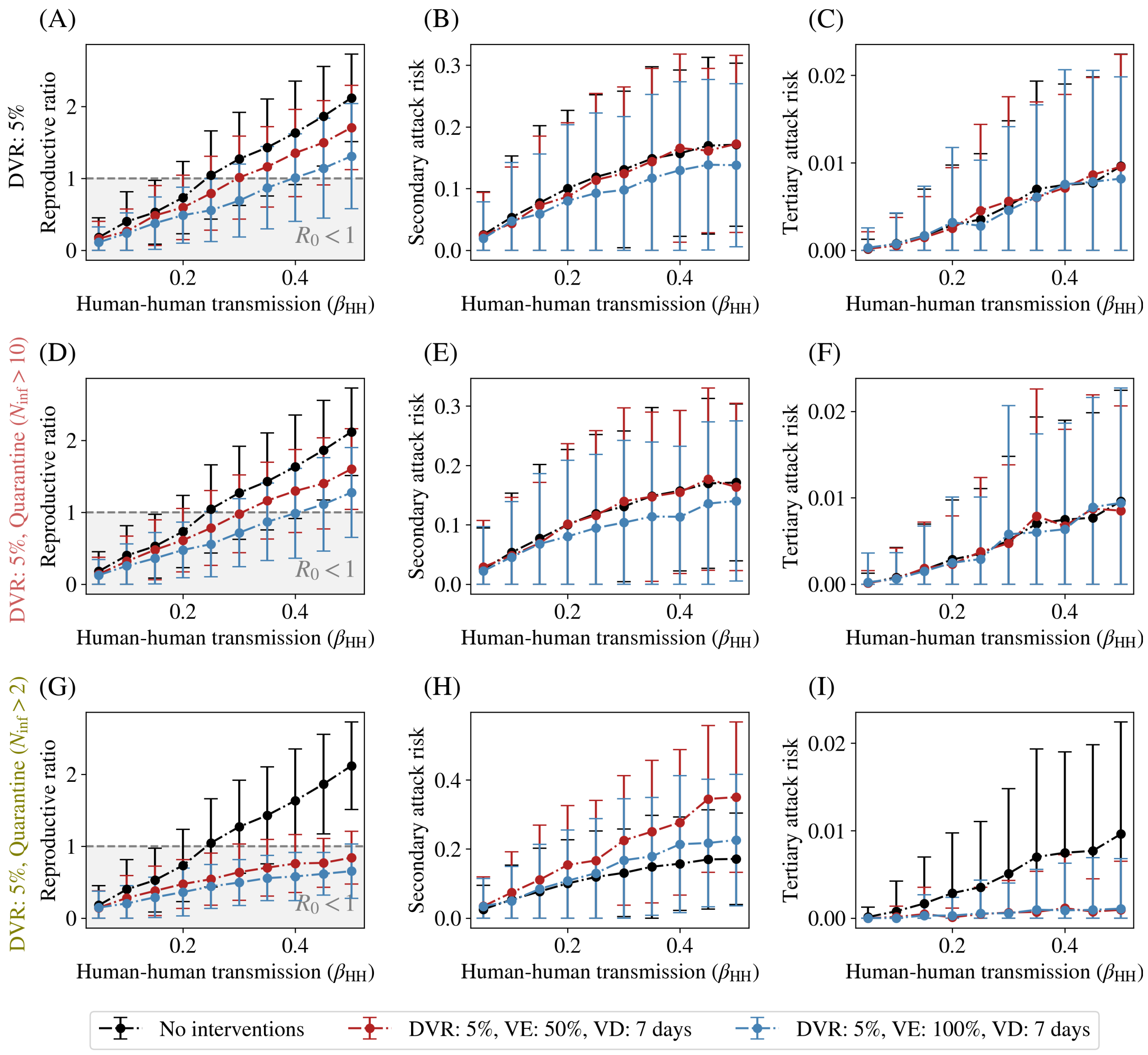


**Fig S4.3: Daily vaccination rate of 5% and delay of 1 week.** We show the same graph as Fig. S4.2, but with a higher daily vaccination rate of 5%. We see that this higher daily vaccination rate causes mean value of the TAR to reduce, but also introduces a lot of variability, which we attribute to many runs having a TAR of 0 due to stochastic effects in which secondary contacts are vaccinated before they get a chance to spread the disease to tertiary contacts. As in the main paper, the error bars in all plots represent the variation (one standard deviation) across 500 different stochastic runs.

#### 4.3 Vaccine delay of 2 weeks

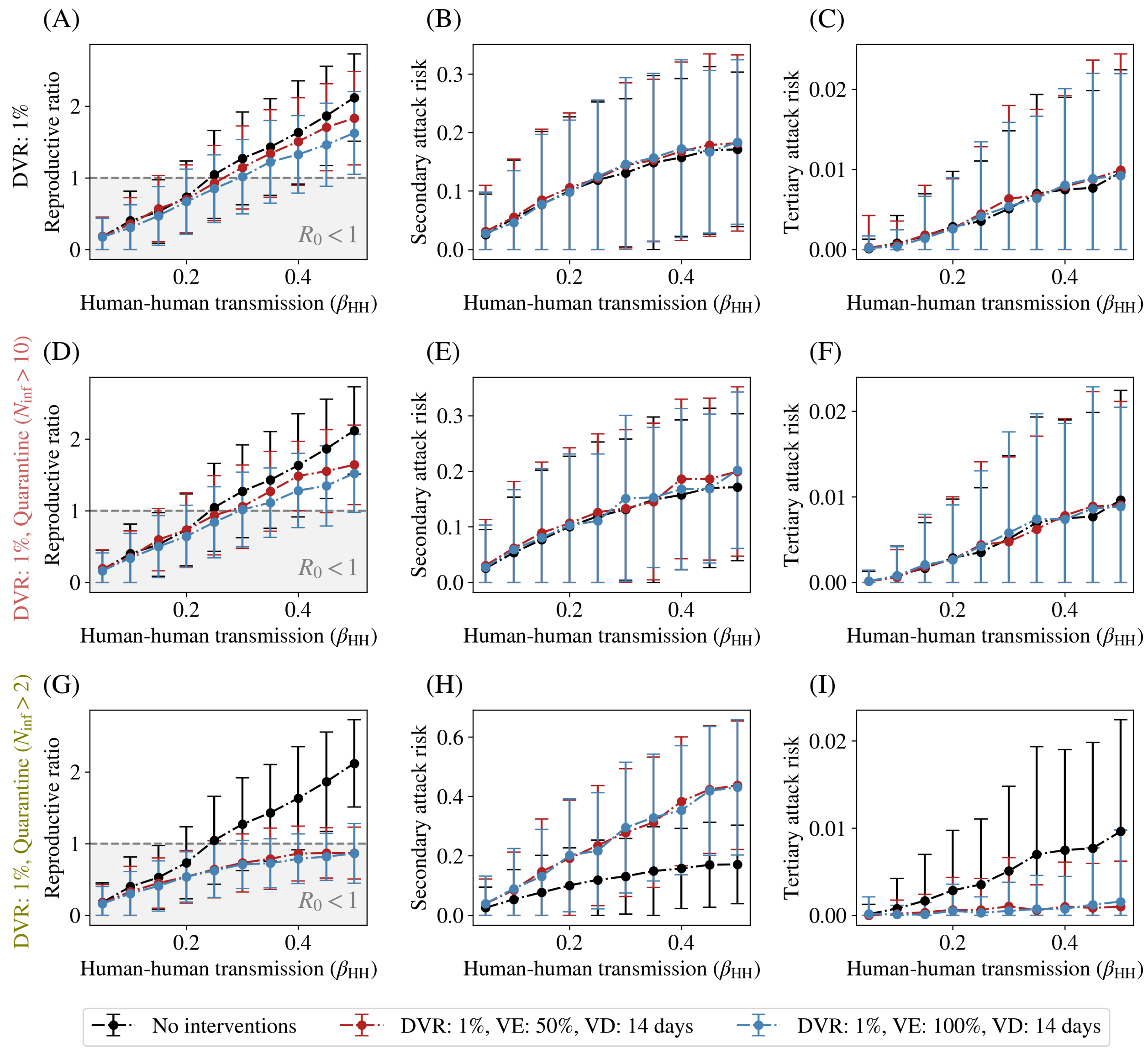


**Fig S4.4: Daily vaccination rate of 1% and delay of 2 weeks.** We now consider a vaccination drive that begins 14 days after the first infectious case has been detected. Here there is a higher probability of the disease spreading to the rest of the population, as evidenced by the high tertiary attack risk. We further see that the efficacy of the vaccine is washed out, especially at higher transmission rates $\beta_{\mathrm{HH}}$. In panels (D), (E), and (F) show the same curves when the primary and secondary contacts are quarantined when 10 infected agents are detected, while panels (G), (H), and (I) show similar graphs, but for a threshold number of 2 infected agents. As in the main paper, the error bars in all plots represent the variation (one standard deviation) across 500 different stochastic runs.

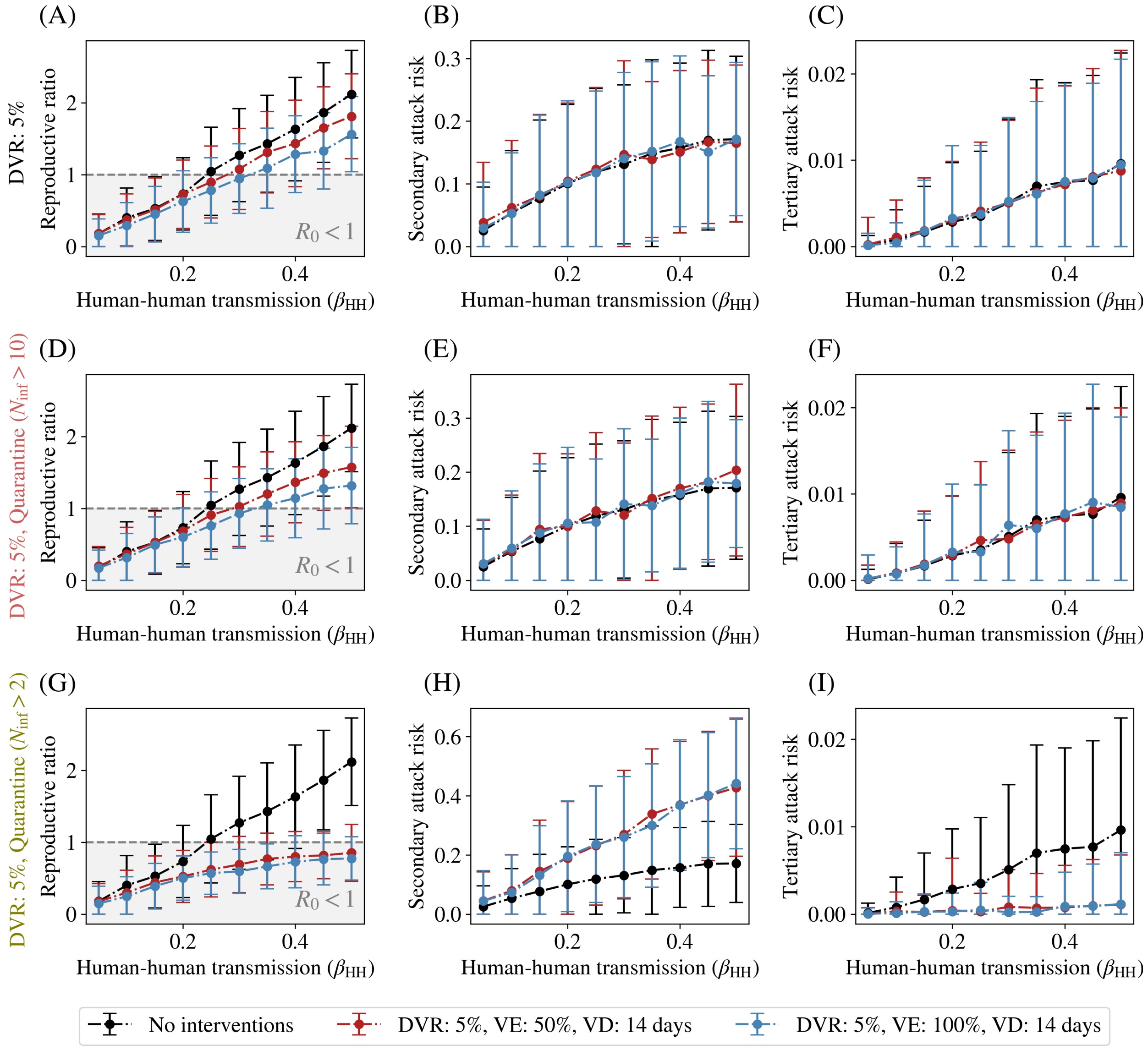


**Fig S4.5: Daily vaccination rate of 5% and delay of 2 weeks.** We show the same graph as Fig. S4.4, but with a higher daily vaccination rate of 5%. We find these results to be very similar to the case of a 1% daily vaccination rate, meaning that with a two week delay, there is very little benefit to having a higher vaccination rate, or vaccine efficacy. This implies that for vaccination to be effective in reducing the spread of the disease, it must be deployed very early in the epidemic. As in the main paper, the error bars in the plots represent the variation (one standard deviation) across 500 different stochastic runs.

### S5 Appendix: Comparing different forces of infection

We consider two different forces of infection: the first based on a well-mixed bird population as described in the paper, and the second using a uniform force of infection. This might represent, for example, a live bird market, where the number of infected birds and other sources of infection are roughly a constant due to steady inflow and outflow.

To compare our results to those in the case of the farm-based outbreak model, the constant force of infection is set by ensuring that it has the same integral as the force of infection derived from the fraction of infected birds.

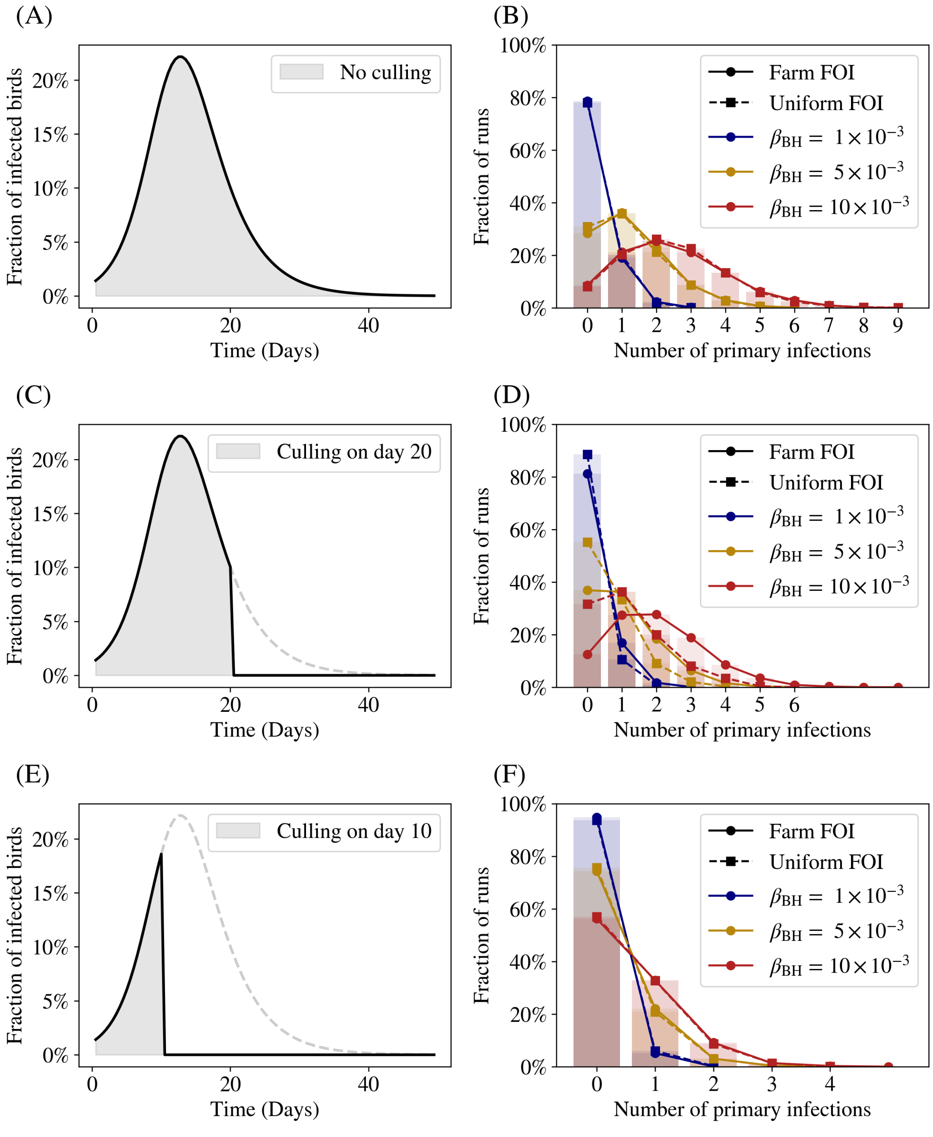


**Fig S5.1: Comparing the distribution of primary cases for different FOIs.** The results of the uniform distribution agree almost exactly with the results from the farm-outbreak FOI for both very early and late culling. However, for intermediate culling, at higher $\beta_{BH}$, clear deviation is observed.

We can now derive the exact form of the distribution of primary cases in both scenarios. We consider first the case of a single primary contact, and compute their survival probability – i.e., the probability that they have *not* been infected by some time $t$ – which we denote $S(t)$. At any given instant of time $t$, the primary contact experiences a hazard function of infection from the birds of

$$p(t) = \begin{cases} \mathcal{F}(t), & 0 < t < 12 \text{ hrs}, \\ 0, & 12 < t < 24 \text{ hrs}, \end{cases} \tag{1}$$

since the primary contacts only spend 12 hours a day in the outbreak site.

From this, we can compute the total risk of infection that the primary contact accumulates up to time $T$, the *cumulative hazard function* $\Lambda(T)$. This quantity can be computed in a mechanistic probabilistic way as follows: we divide time into $n$ small intervals of length $\Delta t = T/n$. At each time-step $i$, the primary contact has a probability $p(t_i)\Delta t$ of being infected. Thus, the probability that the primary contact escapes infection up to some time $t_n$ is just $\Lambda_n$,

$$\Lambda_n = \prod_{i=0}^{n-1} \left(1 - p(t_i)\Delta t\right). \tag{2}$$

In the limit that $\Delta t \to 0$ and $n \to \infty$, we have

$$\Lambda_{n\to\infty,\Delta t\to 0} = \prod_{i=0}^{n-1} (1 - p(t_i)\Delta t) = e^{\sum_{i=0}^{n-1} p(t_k)\Delta t} \to \exp\left(-\int_0^T p(t)\mathrm{d}t\right). \tag{3}$$

Thus, the cumulative hazard function is just

$$\Lambda(T) = \int_0^T p(t)\mathrm{d}t. \tag{4}$$

We can now model this system as a non-homogeneous Poisson process with this hazard function. In such a case, the probability of escaping infection is just the probability of "survival" in the earlier case. Thus, the probability of not being infected is

$$P_{\text{not infected}}(T) = e^{-\Lambda(T)} \qquad \Longrightarrow \qquad P_{\text{infected}}(T) = 1 - e^{-\Lambda(T)}. \tag{5}$$

Thus, every primary contact has an (independent) probability of $P_{\text{infected}}(T) = 1 - e^{-\Lambda(T)}$ of getting infected by time $T$.

We can now move to the case where there are $N$ primary contacts. Since these events are independent, the number of infected primary contacts must form a binomial distribution with a mean of $P_{\text{infected}}(T)$. Thus, the probability that there are exactly $k$ infected primary contacts is

$$P(k \text{ infected primary contacts}) \sim \text{Binomial}\left(N, 1 - e^{-\Lambda(T)}\right) = {}^{N}C_k \left(1 - e^{-\Lambda(T)}\right)^k \left(e^{-\Lambda(T)}\right)^{N-k}. \tag{6}$$

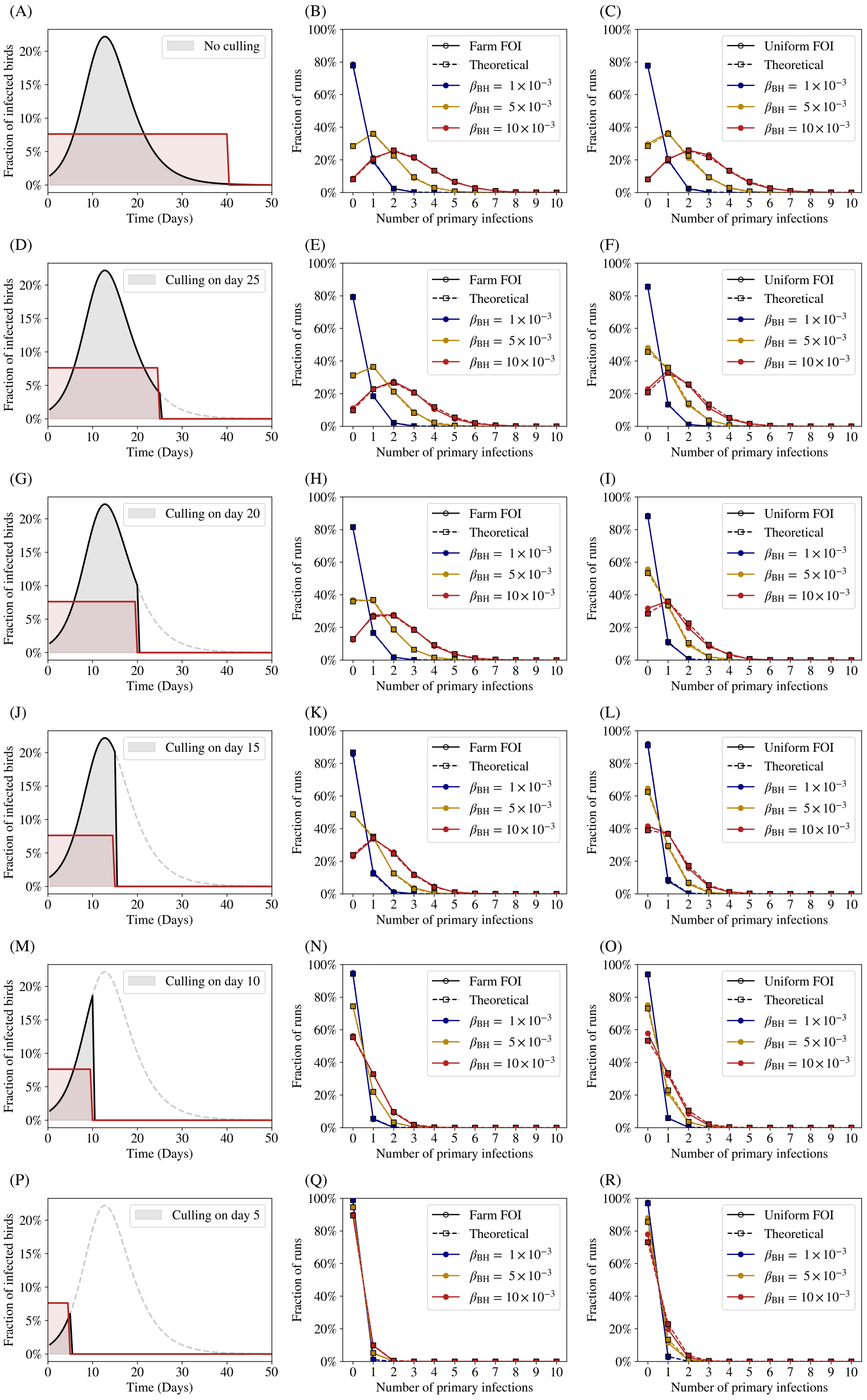


**Fig S5.2: Comparing the distribution of primary cases for different FOIs.** A larger number of CDs is shown, along with a comparison with theoretical results.

### S6 Appendix: Epidemic curves with different interventions

In Fig. S6.1, the total number of recovered humans in the population is averaged over all runs with at least one spillover event and plotted as a function of time for three different scenarios. For simplicity, we restrict ourselves to parameter values that correspond to $R_0 > 1$. As discussed in the main text, quarantining households of primary contacts is the most efficient way of curtailing spread, provided it is done very early.

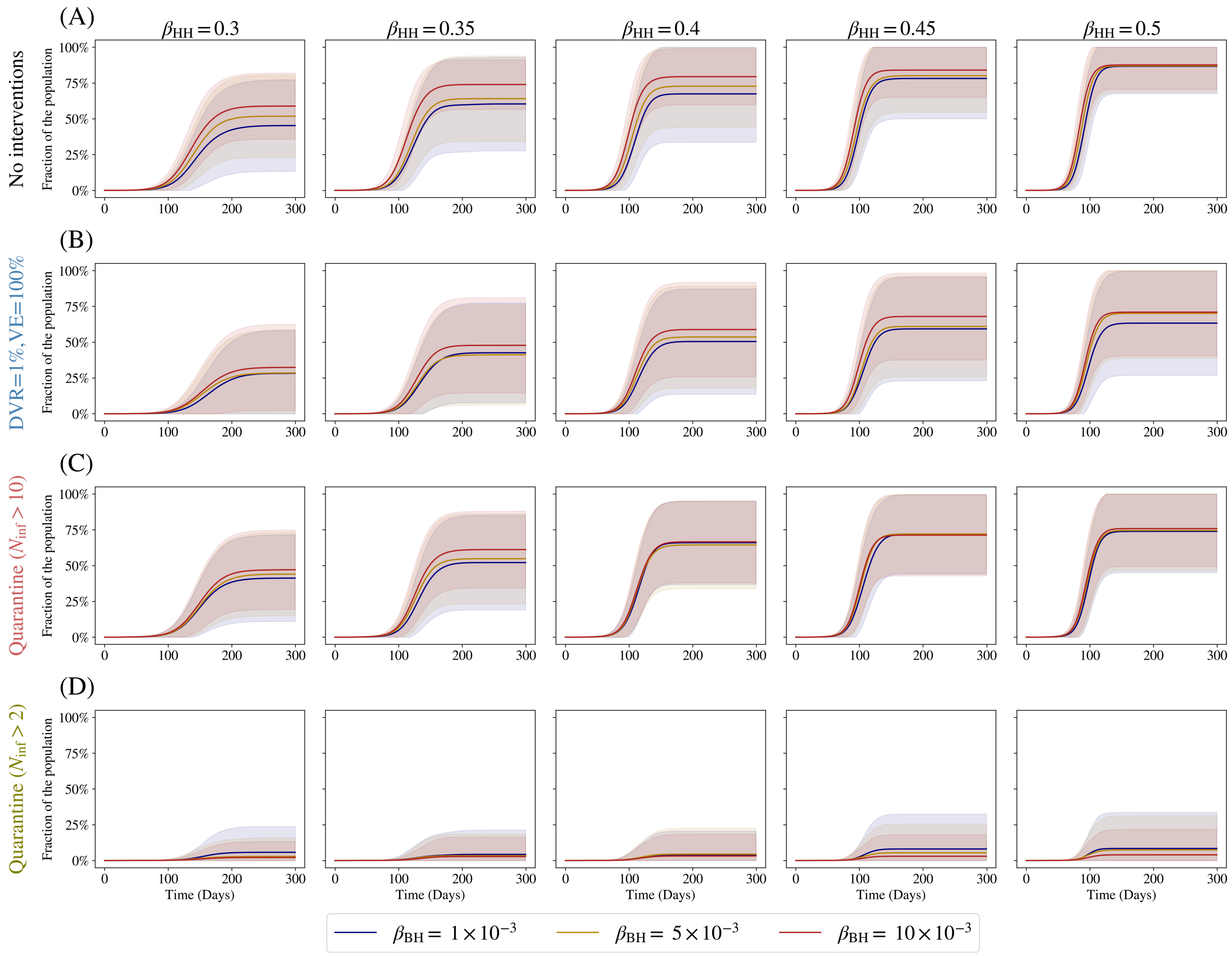


**Fig S6.1: Comparing epidemic curves for different intervention strategies.** In each panel, the different curves represent different values of $\beta_{BH}$. (A) represents the case of no interventions. (B) represents the case where primary and secondary cases are targeted by a vaccination drive which reduces their susceptibility to the disease. The daily vaccination rate (DVR) is set to 1% of the total population, and the vaccine is assumed to be 100% efficient. The vaccine drive begins 1 week after the first infectious case is detected. (C) and (D) represent the cases of a quarantine of primary and secondary cases that occurs after 2 and 10 infectious cases are detected, respectively. We see that quarantining the households of primary and secondary contacts is an efficient way to reduce the spread, provided it is caught very early. The runs in all cases are averages over 500 simulations, with the error-bars being given by $1\sigma$ confidence intervals.

### S7 Appendix: Sensitivity analysis for epidemiological parameters

To see if our results are robust against modest variations in the epidemiological parameters for the disease in humans, we run simulations using an incubation-period ($\tau_E$) and an infectious period ($\tau_I$) in humans that is (i) ∼25% lower and (ii) ∼25% higher than the values used in the main paper, which translates to effectively reducing and increasing the serial interval respectively. In Figs. S7.1 and S7.2, we show the results for vaccination and quarantining, taken separately.

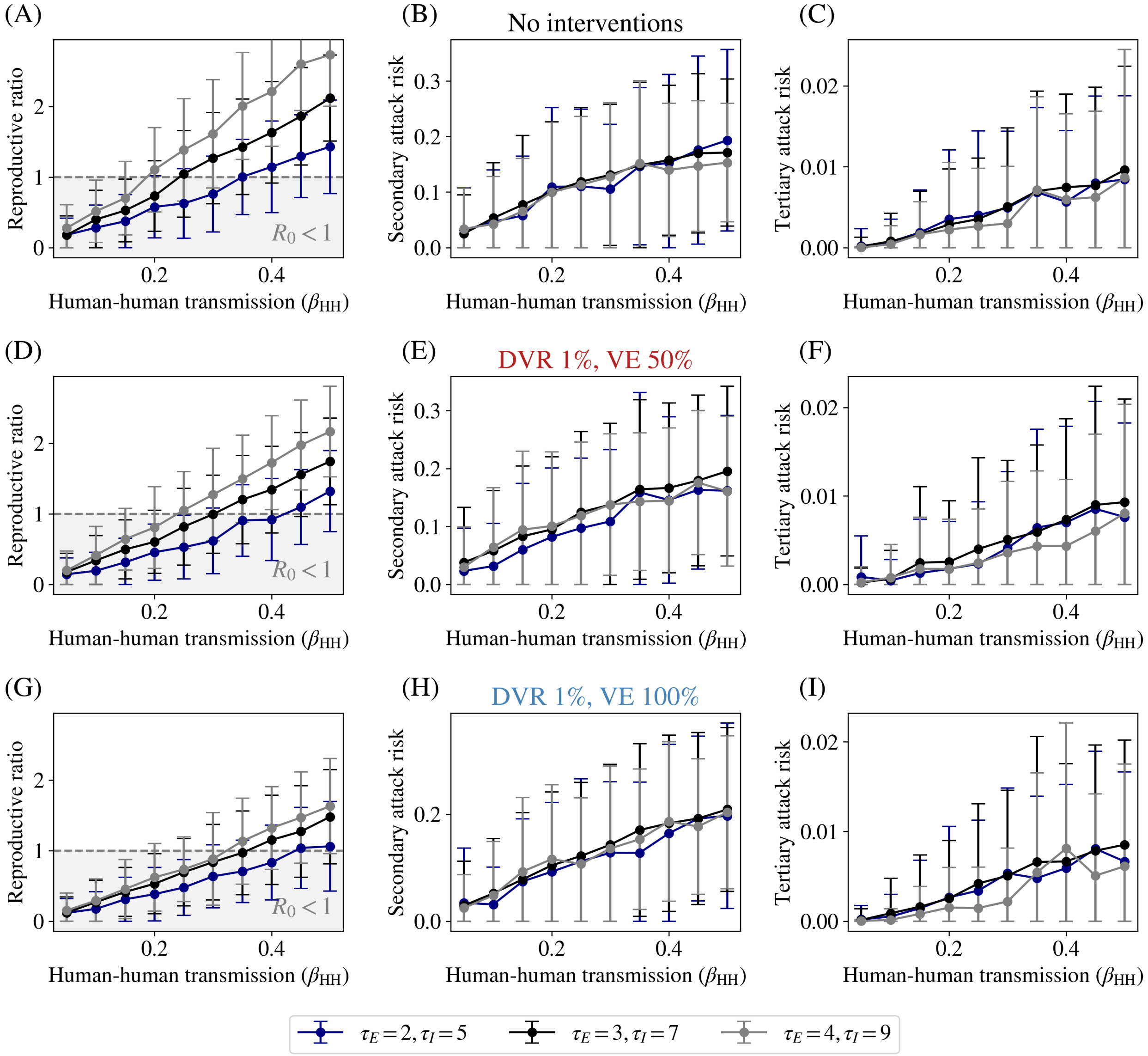


**Fig S7.1: Sensitivity of incubation and infectious periods with vaccination.** Results using different values of the incubation period ($\tau_E$) and infectious period ($\tau_I$) for the case with a 1% daily vaccination rate and a vaccine delay of 1 week. The black curves represent the scenarios from the main paper ($\tau_E = 3, \tau_I = 7$). As can be seen, the reproductive ratio remains linear in $\beta_{\rm HH}$, but depends more (less) steeply on it with increased (decreased) serial interval, but the SAR and TAR are unaffected. As before, the error bars in all plots represent the variation (one standard deviation) across 500 different stochastic runs.

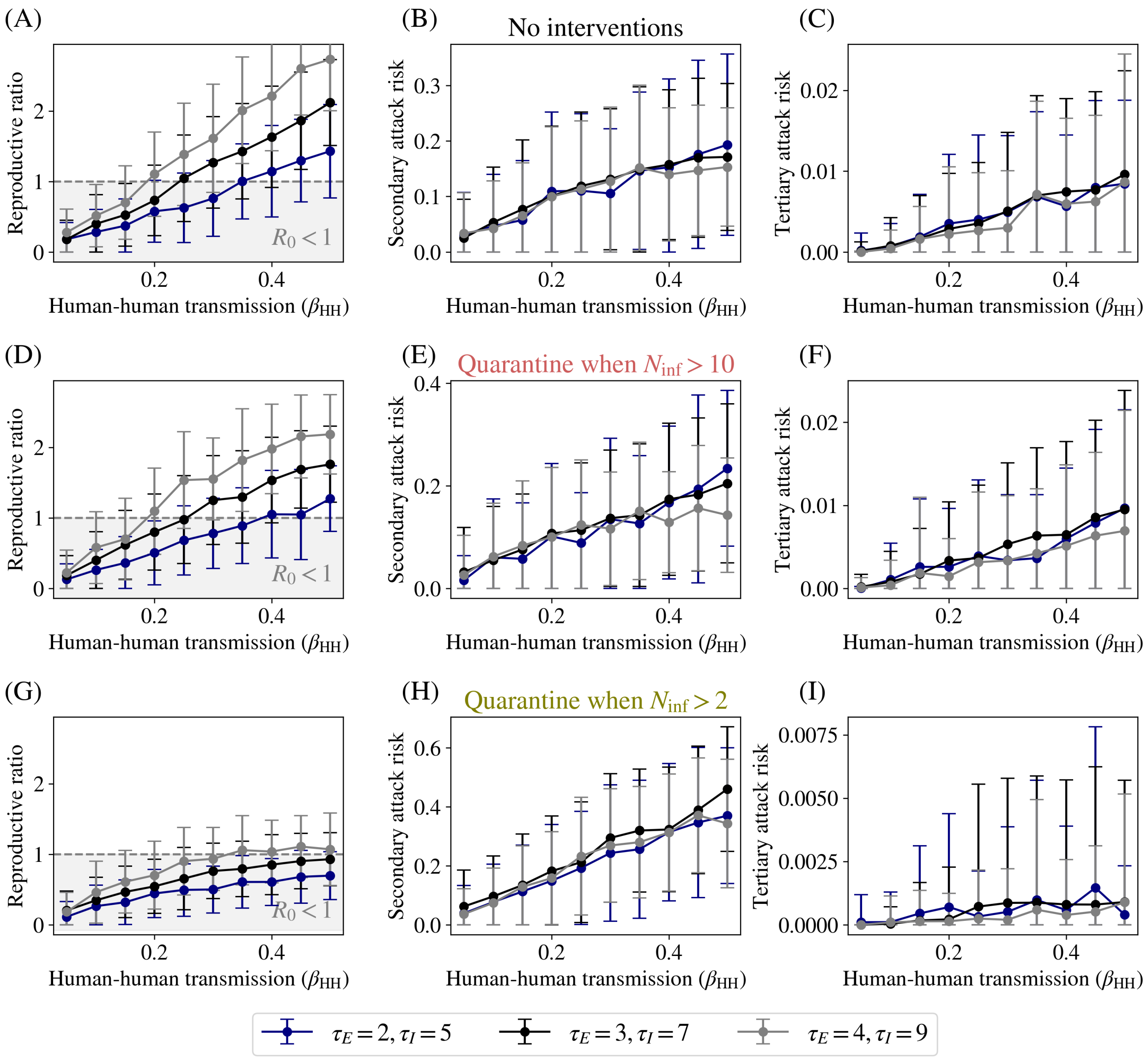


**Fig S7.2: Sensitivity of incubation and infectious periods with quarantine.** Quarantining strategies are qualitatively identical to the case in the main paper (black curves), with the above-mentioned increase in $R_0$ with the serial interval for a given $\beta_{\text{HH}}$. As before, the error bars in all plots represent the variation (one standard deviation) across 500 different stochastic runs.

In each figure, the first row of panels represents the same "no-interventions" scenario. In the case of vaccinations we consider a daily vaccination rate of 1%, and a vaccine delay of 1 week before the drive is begun. Each panel compares the results of a different vaccine efficacy (50% and 100%). Similarly, in the case of quarantining, we consider the two scenarios discussed in the main paper: a quarantine imposed when 10 infected individuals are detected in the population, and one imposed when 2 infected individuals are detected in the population.

Broadly, we find that increasing the serial interval increases the reproductive ratio of the disease (as is to be expected since a longer infectious duration allows for more individuals to be infected) but does not significantly affect the SAR or TAR.
